# Causes and consequences of child growth faltering in low- and middle-income countries

**DOI:** 10.1101/2020.06.09.20127100

**Authors:** Andrew Mertens, Jade Benjamin-Chung, John M Colford, Jeremy Coyle, Mark J van der Laan, Alan E Hubbard, Sonali Rosete, Ivana Malenica, Nima Hejazi, Oleg Sofrygin, Wilson Cai, Haodong Li, Anna Nguyen, Nolan N Pokpongkiat, Stephanie Djajadi, Anmol Seth, Esther Jung, Esther O Chung, Wendy Jilek, Vishak Subramoney, Ryan Hafen, Jonas Häggström, Thea Norman, Kenneth H Brown, Parul Christian, Benjamin F. Arnold, members of the ki Child Growth Consortium

## Abstract

Growth faltering (low length-for-age or weight-for-length) in the first 1000 days — from conception to two years of age — influences short and long-term health and survival. Interventions such as nutritional supplementation during pregnancy and the postnatal period could help prevent growth faltering, but programmatic action has been insufficient to eliminate the high burden of stunting and wasting in low- and middle-income countries. Future preventive efforts will benefit from understanding age-windows and population subgroups in which to focus. Here, we use a population intervention effects analysis of 33 longitudinal cohorts (83,671 children, 662,763 measurements) and 30 separate exposures to show that improving maternal anthropometry and child condition at birth accounted for population increases in length-for-age Z of up to 0.40 and weight-for-length Z of up to 0.15 by age 24 months. Boys had consistently higher risk of all forms of growth faltering than girls. Early post-natal growth faltering predisposed children to subsequent and persistent growth faltering. Children with multiple growth deficits had higher mortality rates from birth to two years than those without deficits (hazard ratios 1.9 to 8.7). The importance of prenatal causes, and severe consequences for children who experienced early growth faltering, support a focus on pre-conception and pregnancy as key opportunities for new preventive interventions.

## Introduction

Child growth faltering in the form of stunting, a marker of chronic malnutrition, and wasting, a marker of acute malnutrition, is common among young children in low-resource settings, and may contribute to child mortality and adult morbidity.^1, 2^ Worldwide, 22% of children under age 5 years are stunted and 7% are wasted, with most of the burden occurring in low- and middle-income counties (LMIC).^3^ Current estimates attribute >250,000 deaths annually to stunting and >1 million deaths annually to wasting.^2^ Stunted or wasted children also experience worse cognitive development^4–6^ and adult economic outcomes.^7^

Despite widespread recognition of the importance of growth faltering to global public health, preventive interventions in LMICs have had limited success.^8^ A range of nutritional interventions targeting various life stages of the fetal and childhood periods, including nutrition education, food and micronutrient supplementation during pregnancy, promotion of exclusive breastfeeding for 6 months and continued breastfeeding for 2 years, and food and micronutrient supplementation during complementary feeding, have all had a beneficial effect on child growth.^9–11^ However, postnatal breastfeeding interventions and nutritional interventions delivered to children who have begun complementary feeding have only had small effects on population-level stunting and wasting burdens, and implementation remains a substantial challenge.^9, 12, 13^ Additionally, water, sanitation, and hygiene (WASH) interventions, which aim to reduce childhood infections that may heighten the risk of wasting and stunting, have had no effect on child growth in several recent large randomized control trials.^14–16^

Modest effects of interventions to prevent stunting and wasting may reflect an incomplete understanding of the optimal way and time to intervene.^17^ This knowledge gap has spurred renewed interest in recent decades to combine rich data sources with advances in statistical methodology^18^ to more deeply understand the key causes of growth faltering.^19^ Understanding the relationship between the causes and timing of growth faltering is also crucial because children who falter early could be at higher risk for more severe growth faltering later. In companion articles, we report that the highest rates of incident stunting and wasting occur by age 3 months.^20, 21^

### Pooled longitudinal analyses

Here, we report a pooled analysis of 33 longitudinal cohorts in 15 low- and middle-income countries in South Asia, Sub-Saharan Africa, Latin America, and Eastern Europe, with data collection initiated between 1987 and 2014. Our objective was to estimate relationships between child, parental, and household characteristics and measures of child anthropometry, including length-for-age Z-scores (LAZ), weight-for-length Z-scores (WLZ), weight-for-age Z-scores (WAZ), stunting, wasting, underweight, and length and weight velocities from birth to age 24 months. Details on the estimation of growth faltering outcomes are included in companion articles.^20, 21^ We also estimated associations between early growth faltering and more severe growth faltering or mortality by age 24 months.

Cohorts were assembled as part of the Bill & Melinda Gates Foundation’s Knowledge Integration (*ki*) initiative, which included studies of growth and development in the first 1000 days, beginning at conception.^22^ We selected longitudinal cohorts from the database that met five inclusion criteria: 1) conducted in low- or middle-income countries; 2) enrolled children between birth and age 24 months and measured their length and weight repeatedly over time; 3) did not restrict enrollment to acutely ill children; 4) enrolled children with a median year of birth after 1990; and 5) collected anthropometric status measurements at least every 3 months (Extended Data Fig 1). Inclusion criteria ensured we could rigorously evaluate the timing and onset of growth faltering among children who were broadly representative of populations in low- and middle-income countries. Thirty-three cohorts from 15 countries met inclusion criteria, and 83,671 children and 592,030 total measurements were included in this analysis (Fig 1). Child mortality was rare and not reported in many of the *ki* datasets, so we relaxed inclusion criteria for studies used in the mortality analysis to include studies that measured children at least twice a year. Four additional cohorts met this inclusion criterion, and 14,317 children and 70,733 additional measurements were included in mortality analyses (97,988 total children, 662,763 total observations, Extended Data Table 1). Cohorts were distributed throughout South Asia, Africa, and Latin America, with a single European cohort from Belarus.

**Figure 1.**
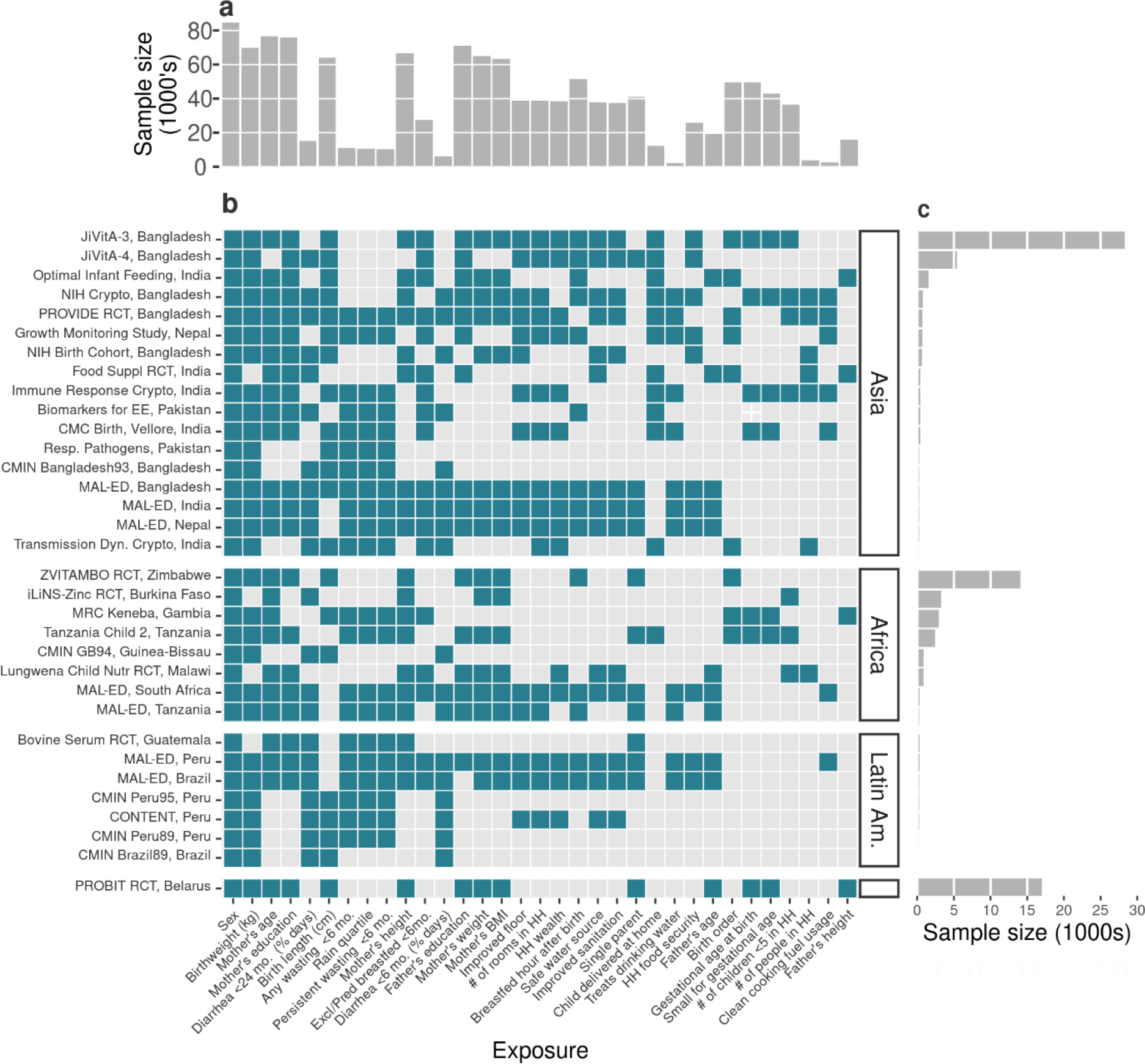
Cohort sample sizes and exposures measured. (**a**) Total number of children with each measured exposure, sorted from left to right by number of cohorts measuring the exposure. (**b**) Presence of 30 exposure variables in the *ki* data by within each included cohort. Cohorts are sorted by geographic region and sample size. (**c**) Number child anthropometry observations contributed by each cohort.

### Population intervention effects on growth faltering

In a series of analyses, we estimated population intervention effects, the estimated change in population mean Z-score if all individuals in the population had their exposure shifted from observed levels to the lowest-risk reference level.^23^ The PIE is a policy-relevant parameter; it estimates the improvement in outcome that could be achievable through intervention for modifiable exposures, as it is a function of the degree of difference between the unexposed and the exposed in a children’s anthropometry Z-scores, as well as the observed distribution of exposure in the population. We selected exposures that were measured in multiple cohorts, could be harmonized across cohorts for pooled analyses, and had been identified as important predictors of stunting or wasting in prior literature (Fig 1, Extended Data Table 2). Exposure measurement varied by cohort, but all estimates were adjusted for all other measured exposures that we assumed were not on the causal pathway between the exposure of interest and the outcome. For example, the association between maternal height and stunting was not adjusted for child birth weight, because low maternal height could increase stunting risk through lower child birth weight.^24^ Parameters were estimated using targeted maximum likelihood estimation, a doubly-robust, semiparametric method that allows for valid inference while adjusting for potential confounders using ensemble machine learning (details in Methods).^18, 25^ We estimated cohort-specific parameters, adjusting for measured covariates within each cohort, and then pooled estimates across cohorts using random effects models (Extended Data Fig 1).^26^ We chose as the reference exposure for PIEs the level of lowest risk across cohorts. We also estimated the effects of optimal dynamic interventions, where each child’s individual low-risk level of exposure was estimated from potential confounders (details in Methods). Timing of exposures varied, from parental and household characteristics present before birth, to fetal or at-birth exposures, and postnatal exposures. We estimated associations with growth faltering that occurred after exposure measurements to ensure time-ordering of exposures and outcomes.

Population level improvements in maternal height and birth size would be expected to improve child LAZ and WLZ at age 24 months substantially, owing to both the high prevalence of suboptimal anthropometry in the populations and their strong association with attained growth at 24 months (Fig 2, Fig 3). Beyond anthropometry, key predictors of higher Z-scores included markers of better household socioeconomic status (e.g., number of rooms in the home, parental education, clean cooking fuel use, household wealth index). The pooled, cross-validated R^2^ for models that included the top 10 determinants for each Z-score, plus child sex, was 0.25 for LAZ (N= 20 cohorts, 25,647 children) and 0.07 for WLZ (N=18 cohorts, 17,853 children). The population level impact of season on WLZ was large, with higher WLZ in drier periods (Fig 3), consistent with seasonal differences shown in a companion article.^21^ Exclusive or predominant breastfeeding before 6 months of age was associated with higher WLZ but not LAZ at 6 months of age and was not a major predictor of Z-scores at 24 months (Extended Data Figs 2,3,4).^27^ Girls had consistently better LAZ and WLZ than boys, potentially from sex-specific differences in immunology, nutritional demands, care practices, and intrauterine growth.^28^

**Figure 2.**
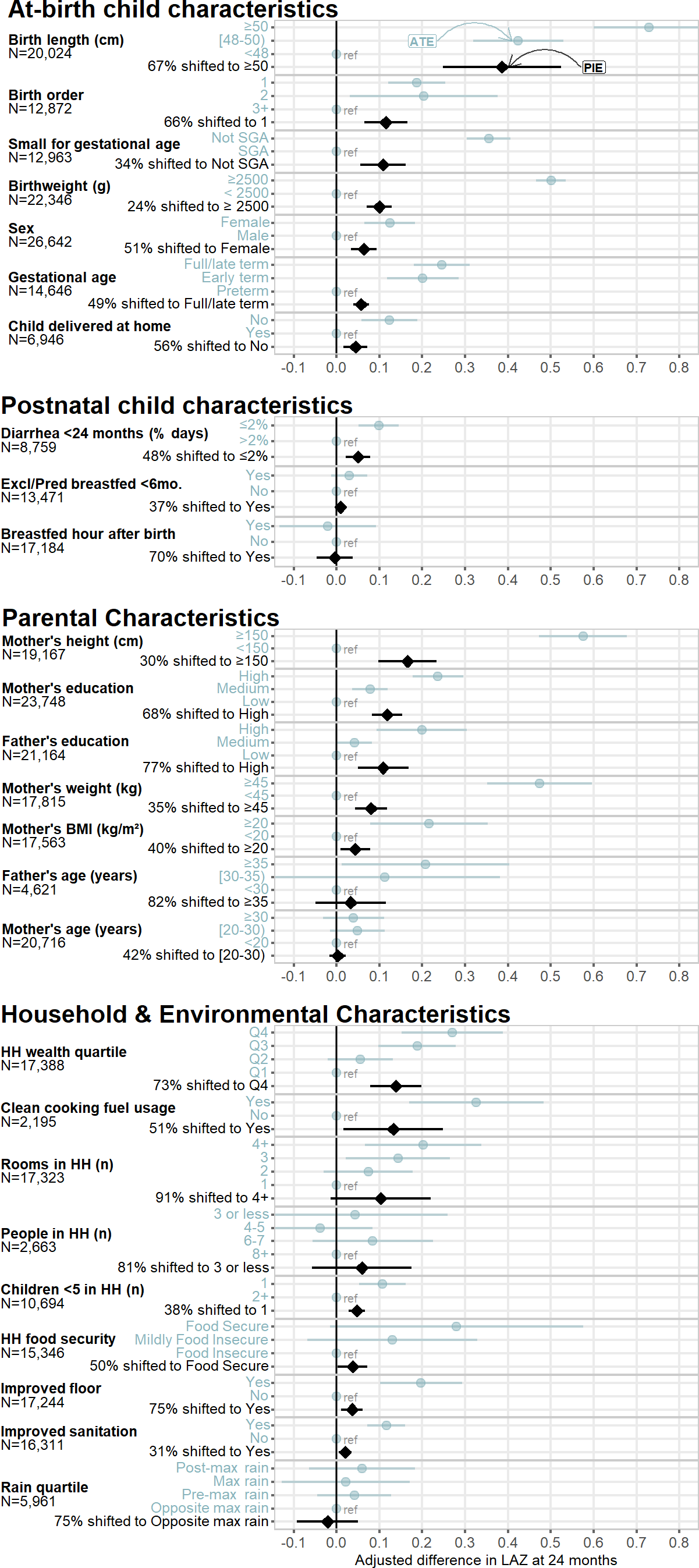
Population intervention effects and mean differences of child, parental, and household exposures on length-for-age z-scores at age 24 months. Lighter points show adjusted mean differences (average treatment effects, ATEs) between the labeled higher-risk levels of exposures and reference levels, and black points show population intervention effects (PIEs), the estimated difference in length-for-age z-scores (LAZ) after shifting exposure levels for all children to the reference level. The number of children that contributed to each analysis is listed by exposure. The colored Y-axis label is either the level of exposure contrasted against the reference level to estimate the ATE, or the percentage of the population shifted to the lowest-risk level to estimate the PIE. Cohort-specific estimates were adjusted for all measured confounders using ensemble machine learning and TMLE, and then pooled using random effects (Methods). Estimates are shown only for exposures measured in at least 4 cohorts.

**Figure 3.**
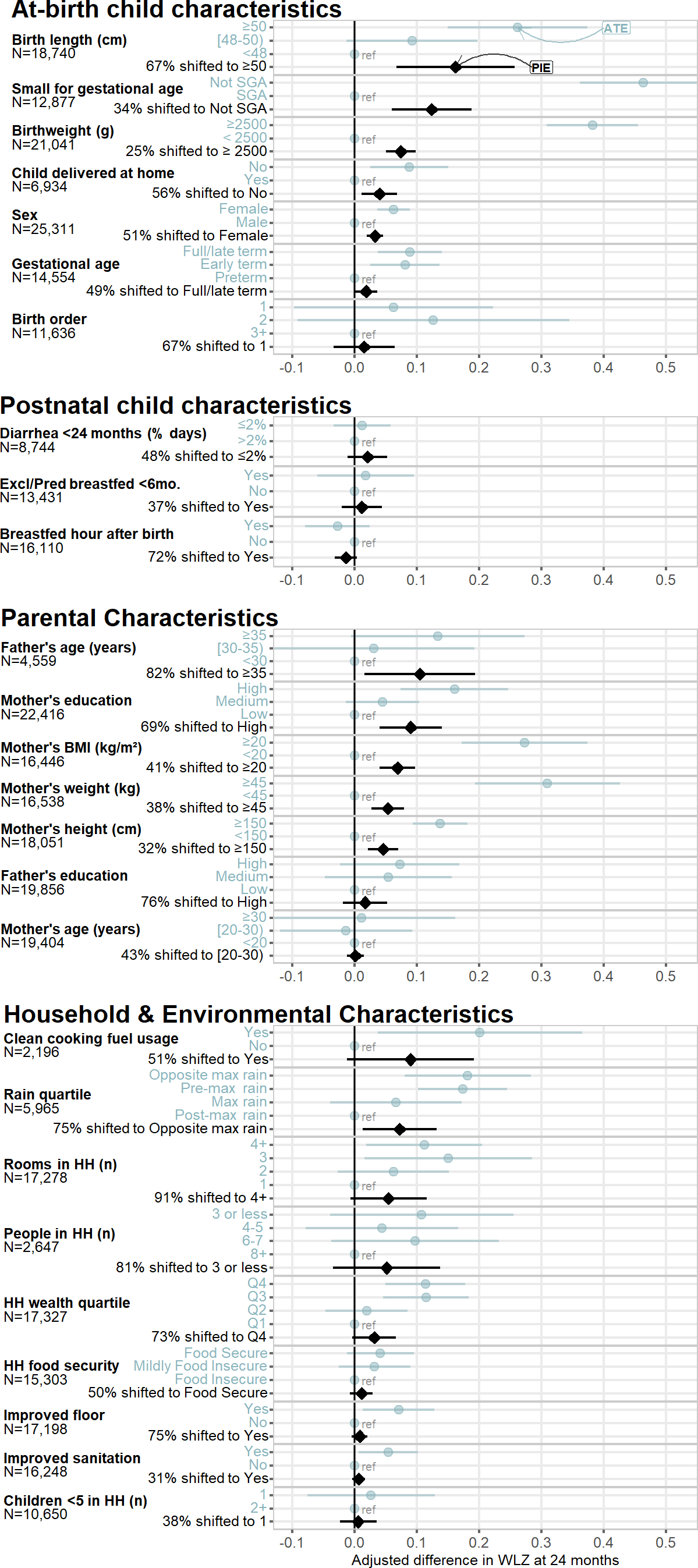
Population intervention effects and mean differences of child, parental, and household exposures on weight-for-length z-scores at age 24 months. Lighter points show adjusted mean differences (average treatment effects, ATEs), between higher-risk exposure levels and reference levels, and black points show population intervention effects (PIEs), the estimated difference in weight-for-length z-scores (WLZ) after shifting exposure levels for all children to the reference level. The number of children that contributed to each analysis is listed by exposure. The colored Y-axis label is either the level of exposure contrasted against the reference level to estimate the ATE, or the percentage of the population shifted to the lowest-risk level to estimate the PIE. Cohort- specific estimates were adjusted for all measured confounders using ensemble machine learning and TMLE, and then pooled using random effects (Methods). Estimates are shown only for exposures measured in at least 4 cohorts.

The findings underscore the importance of prenatal exposures for child growth outcomes, and at the population-level growth faltering may be difficult to shift without broad improvements in standard of living.^7, 29^ Maternal anthropometric status can influence child Z-scores by affecting fetal growth and birth size.^30, 31^ Maternal height and BMI could directly affect postnatal growth through breastmilk quality, or could reflect family poverty, genetics, undernutrition, or food insecurity, or family lifestyle and diet.^32, 33^ In a secondary analysis, we estimated the associations between parental anthropometry and child Z-scores controlling for birth characteristics, and found the associations were only partially mediated by birth size, order, hospital delivery, and gestational age at birth, with adjusted Z-score differences attenuated by a median of 30% (Extended Data Fig 5).

**Figure 5.**
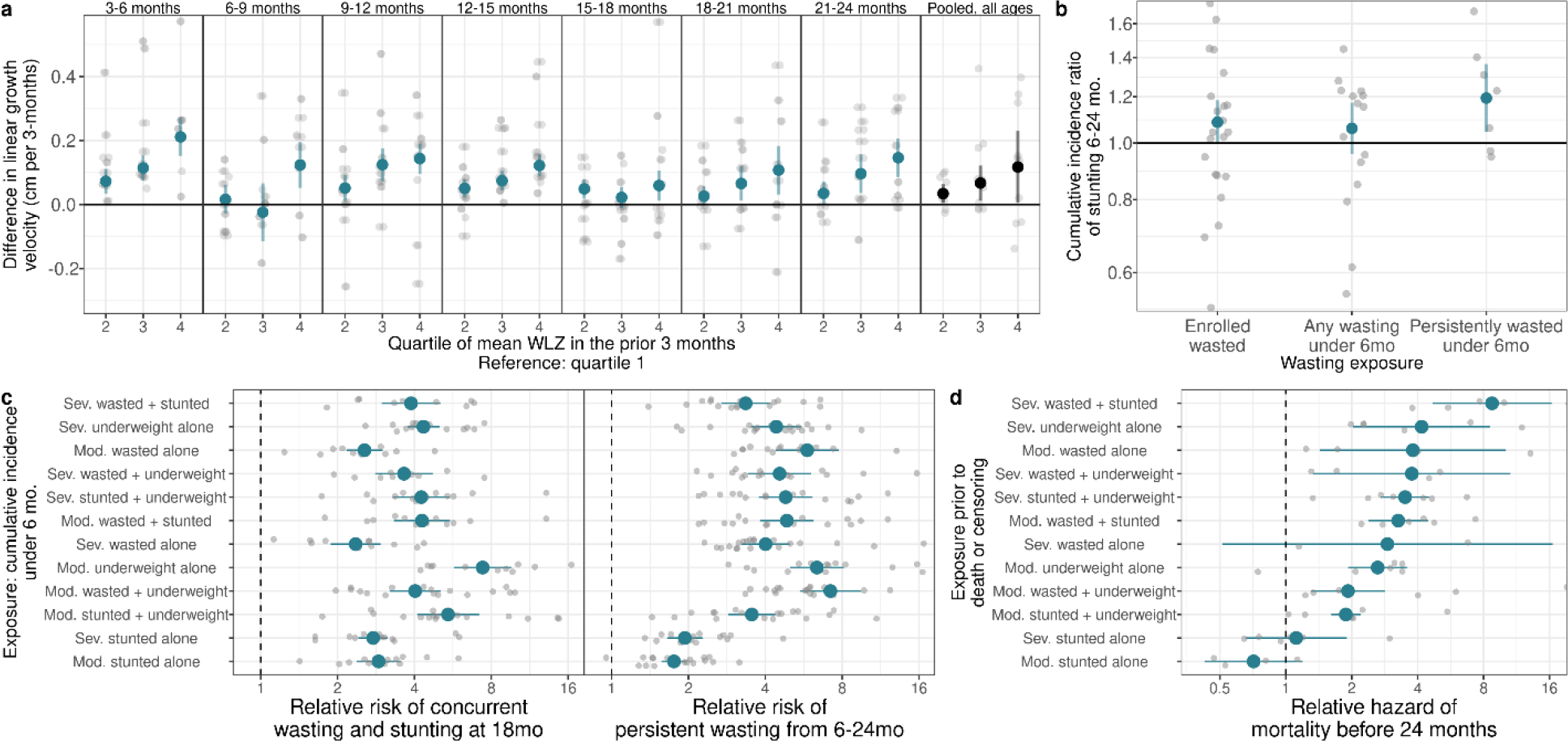
Early life growth faltering increases risk of more severe growth faltering and mortality. (a) Adjusted differences in linear growth velocity (in centimeters) across 3-month age bands, by quartile of weight-for-length z-score (WLZ) in the preceding three months. The reference group is children in the first quartile of WLZ in the previous age period. The panel with black points on the far right shows the pooled estimates, unstratified by child age. Velocity was calculated from the closest measurements within 14 days of the start and end of the age period. (**b**) Relative risk of stunting onset between ages 6-24 months among children who experienced measures of early wasting before age 6 months compared to children who did not. Gray points indicate cohort-specific estimates. (**c**) Association between cumulative incidence of mutually exclusive definitions of growth faltering before age 6 months (reference: children with no measure of growth failure) and persistent wasting from ages 6-24 months (33 cohorts, 6,046 cases, and 68,645 children) or concurrent wasting and stunting at 18 months. (31 cohorts, 1,447 cases, and 22,565 children). Growth faltering definitions are sorted by estimates in panel **d**. (**d**) Hazard ratios between mutually exclusive definitions of growth faltering (reference: children with no measure of growth failure) and mortality before 24 months (8 cohorts, 1,689 deaths with ages of death, and 63,812 children). Gray points indicate cohort-specific estimates.

The strongest predictors of stunting and wasting estimated through population attributable fractions closely matched those identified for child LAZ and WLZ at 24 months (Extended Data Figs 6, 7), suggesting that information embedded in continuous and binary measures of child growth provide similar inference with respect to identifying public-health relevant causes. Potential improvements through population interventions were relatively modest. For example, if all children were born to higher BMI mothers (≥ 20) compared to the observed distribution of maternal BMI, one of the largest predictors of wasting, we estimate it would reduce the incidence of wasting by age 24 months by 8.2% (95% CI: 4.4, 12.0; Extended Data Fig 7). Patterns in associations across growth outcomes were broadly consistent, except for preterm birth, which had a stronger association with stunting outcomes than wasting outcomes, and rainy season, which was strongly associated with wasting but not stunting (Extended Data Fig 2). Direction of associations did not vary across regions, but magnitude did, notably male sex was less strongly associated with low LAZ in South Asia (Extended Data Figs 8,9).

### Age-varying effects on growth faltering

We estimated trajectories of mean LAZ and WLZ stratified by maternal height and BMI. We found that maternal height strongly influenced at-birth LAZ, but that LAZ progressed along similar trajectories through age 24 months regardless of maternal height (Fig 4a), with similar though slightly less pronounced differences when stratified by maternal BMI (Fig 4b). By contrast, children born to taller mothers had similar WLZ at birth and WLZ trajectories until age 3-4 months, when they diverged substantially (Fig 4a); WLZ trajectory differences were even more pronounced when stratified by maternal BMI (Fig 4b). The findings illustrate how maternal status strongly influences where child growth trajectories start, but that growth trajectories evolve in parallel, seeming to respond similarly to postnatal insults independent of their starting point.

**Figure 4.**
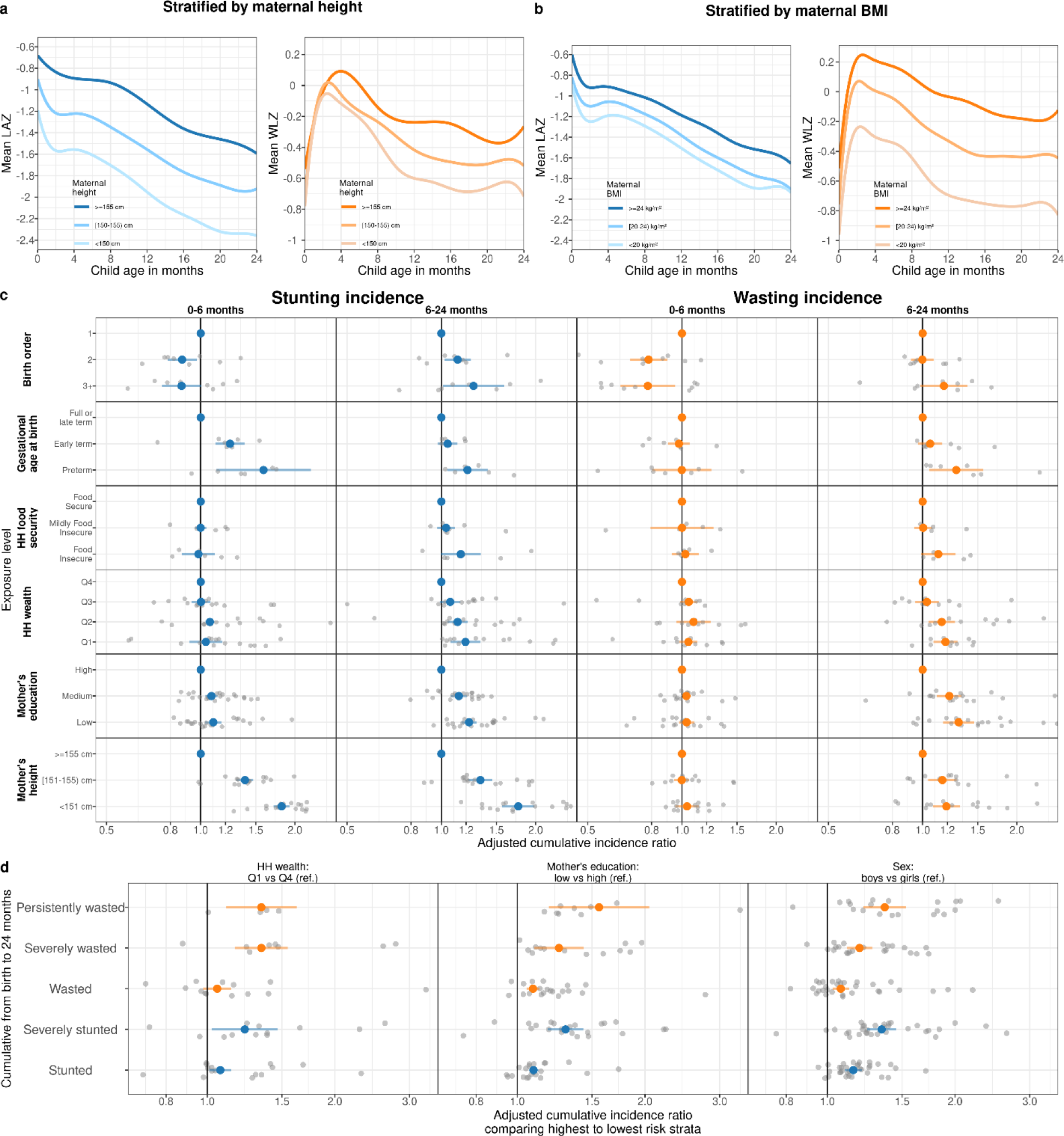
Effect of key exposures on the trajectories, timing, and severity of child **growth faltering** (**a**) Child length-for-age Z-score (LAZ) and weight-for-length Z-score (WLZ) trajectories, stratified by categories of maternal height (N=413,921 measurements, 65,061 children, 20 studies). (**b**) Child LAZ and WLZ, stratified by categories of maternal BMI (N=373,382 measurements, 61,933 children, 17 studies). Growth trajectories stratified by all risk factors examined beyond maternal height and BMI are available online (https://child-growth.github.io/causes/rf-splines.html). (**c**) Associations between key exposures and wasting cumulative incidence, stratified by the age of the child during wasting incidence. Gray points indicate cohort-specific estimates. (**d**) Associations between key exposures and growth faltering of different severity. Cumulative incidence ratios contrast the highest and lowest risk categories of each exposure, which are printed in each panel title. Gray points indicate cohort-specific estimates.

We hypothesized that causes of growth faltering could differ by age of growth faltering onset — for example, we expected children born preterm would have higher risk of incident growth faltering immediately after birth, while food insecurity might increase risk at older ages, after weaning. For exposures studied in the population intervention effect analyses, we conducted analyses stratified by age of onset and in many cases found age-varying effects (Fig 4c). For example, most measures of socioeconomic status were associated with incident wasting or stunting only after age 6 months, and higher birth order lowered growth faltering risk under age 6 months, but increased risk thereafter. First born babies are born thin and show rapid postnatal catch up in WLZ (Extended data Fig 10). This is likely because first-born babies suffer uterine constraint caused by a less developed uterine-placental-vascular supply^34, 35^ resulting in birth weights being lower by 100-200g in most cohorts studied; weight is generally more compromised than height.^36^ The switch from a constrained uterine-placental nutrient supply line to oral nutrition permits post-natal catch up. Stronger relationships between key socio- demographic characteristics and wasting and stunting as children age likely reflects the accumulation of insults that result from household conditions, particularly as complementary feeding is initiated, and children begin exploring their environment and potentially face higher levels of food insecurity especially in homes with multiple children.^37^ When viewed across multiple definitions of growth faltering, most exposures had stronger associations with severe stunting, severe wasting, or persistent wasting (> 50% of measurements < –2 WLZ), rarer but more serious outcomes, than with incidence of any wasting or stunting (Fig 4d). Additionally, the characteristics strongly associated with lower wasting recovery by 90 days (birth size, small maternal stature, lower maternal education, later birth order, and male sex) increased risk of wasting prevalence and cumulative incidence (Extended Data Fig 2).

### Consequences of early growth faltering

We documented high incidence rates of wasting and stunting from birth to age 6 months in companion papers.^20, 21^ Based on previous studies, we hypothesized that early wasting could contribute to subsequent linear growth restriction, and early growth faltering could be consequential for persistent growth faltering and mortality during the first 24 months of life.^38–40^ Among cohorts with monthly measurements, we examined age-stratified linear growth velocity by quartiles of WLZ at previous ages. We found a consistent exposure-response relationship between higher mean WLZ and faster linear growth velocity in the following 3 months (Fig 5a). Persistent wasting from birth to 6 months (defined as >50% of measurements wasted) was the wasting exposure most strongly associated with incident stunting at older ages (Fig 5b).

We next examined the relationship between measures of growth faltering in the first 6 months and serious growth-related outcomes: persistent wasting from 6-24 months and concurrent wasting and stunting at 18 months of age, both of which put children at high risk of mortality.^1, 38^ Concurrent wasting and stunting was measured at 18 months because stunting prevalence peaked at 18 months and the largest number of children were measured at 18 months across cohorts.^20^ All measures of early growth faltering were significantly associated with later, more serious growth faltering, with measures of ponderal growth faltering amongst the strongest predictors (Fig 5c).

Finally, we estimated hazard ratios (HR) of all-cause mortality by 2 years of age associated with measures of growth faltering within eight cohorts that reported ages of death, which included 1,689 child deaths by age 24 months (2.4% of children in the eight cohorts). Included cohorts were highly monitored, and mortality rates were lower than in the general population in most cohorts (Extended Data Table 3). Additionally, data included only deaths that occurred after anthropometry measurements, so many neonatal deaths may have been excluded, and without data on cause-specific mortality, some deaths may have occurred from causes unrelated to growth faltering. Despite these caveats, growth faltering increased the hazard of death before 24 months for all measures except stunting alone, with strongest associations observed for severe wasting, stunting, and underweight (HR=8.7, 95% CI: 4.7 to 16.4) and severe underweight alone (HR=4.2, 95% CI: 2.0 to 8.6) (Fig 5d).

## Discussion

This synthesis of LMIC cohorts during the first 1000 days of life has provided new insights into the principal causes and near-term consequences of growth faltering. Our use of a novel, semi-parametric method to adjust for potential confounding provided a harmonized approach to estimate population intervention effects that spanned child-, parent-, and household-level exposures with unprecedented breadth (30 exposures) and scale (662,763 anthropometric measurements from 33 cohorts). Our focus on effects of shifting population-level exposures on continuous measures of growth faltering reflect a growing appreciation that growth faltering is a continuous process.^41^ Our results show children in LMICs stand to benefit from interventions to support optimal growth in the first 1000 days. Combining information from high-resolution, longitudinal cohorts enabled us to study critically important outcomes not possible in smaller studies or in cross-sectional data, such as persistent wasting and mortality, as well as examine risk-factors by age.

Maternal, prenatal, and at-birth characteristics were the strongest predictors of growth faltering across regions in LMICs. Our results underscore prenatal exposures as key determinants of child growth faltering.^42^ Limited impact of exclusive or predominant breastfeeding through 6 months (+0.01 LAZ) aligns with a meta-analysis of breastfeeding promotion,^27^ but our finding of limited impact of reducing diarrhea through 24 months (+0.05 LAZ) contrasts with some observational studies.^43, 44^ Many predictors, like child sex, birth order, or season, are not modifiable but could guide interventions that mitigate their effects, such as seasonally targeted supplementation or enhanced monitoring among boys. Strong associations between maternal anthropometry and early growth faltering highlight the role of intergenerational transfer of growth deficits between mothers and their children.^32^ Shifting several key population exposures (maternal height or BMI, education, birth length) to their observed low-risk level would improve LAZ by up to 0.40 z and WLZ by up to 0.15 z in target populations and could be expected to prevent 8% to 32% of incident stunting and wasting (Figs 2,3 Extended Data Figs 6,7). Maternal anthropometric status strongly influenced birth size, but the parallel drop in postnatal Z-scores among children born to different maternal phenotypes was much larger than differences at birth, indicating that growth trajectories were not fully “programmed” at birth (Fig 4a-b). This accords with the transition from a placental to oral nutrient supply at birth.

The analyses have caveats. Population intervention effects were based on exposure distributions in the 33 cohorts, which were not necessarily representative of the general population in each setting. Use of external exposure distributions from population-based surveys would be difficult because many key exposures we considered, such as at-birth characteristics or longitudinal diarrhea prevalence, are not measured in such surveys. In some cases, detailed exposure measurements like longitudinal breastfeeding or diarrhea history were coarsened to simpler measures to harmonize definitions across cohorts, potentially attenuating their association with growth faltering. Other key exposures such as dietary diversity, nutrient consumption, micronutrient status, maternal and child morbidity indicators, pathogen-specific infections, and sub-clinical inflammation and intestinal dysfunction were measured in only a few cohorts, so were not included.^45, 46^ The absence of these exposures in the analysis, some of which have been found to be important within individual contributed cohorts,^46, 47^ means that our results emphasize exposures that were more commonly collected, but likely exclude some additional causes of growth faltering. A final caveat is that we studied consequences through age 24 months — the primary age range of contributed *ki* cohort studies — and thus have not considered effects on longer-term outcomes. Several studies have suggested that puberty could be another potential window for intervention to enhance catch-up growth.^48^ Improving girls’ stature at any point through puberty could help blunt the intergenerational transfer of growth faltering by improving maternal height,^49^ which in turn could improve outcomes among their children (Figs 2, 3, 4a, 4b).

Countries that have reduced stunting most have undergone improvements in maternal education, nutrition, reductions in number of pregnancies, and maternal and newborn health care,^50^ reinforcing the importance of interventions during the window from conception to one year, when fetal and infant growth velocity is high and energy expenditure for growth and development is about 50% above adult values (adjusted for fat-free mass).^51^ A stronger focus on prenatal interventions should not distract from renewed efforts for postnatal prevention. The observed pre- and postnatal growth faltering we observed reinforce the need for sustained support of mothers and children throughout the first 1000 days. Efficacy trials that delivered prenatal nutrition supplements to pregnant mothers,^52–54^ therapeutics to reduce infection and inflammation for pregnant mothers,^55–59^ and nutritional supplements to children 6-24 months^11, 60^ have reduced child growth faltering but have fallen short of completely preventing it.

Our results suggest that the next generation of preventive interventions should focus on the early period of a child’s first 1000 days — from preconception through the first months of life —because maternal status and at-birth characteristics are key determinants of growth faltering through 24 months. Halting the cycle of growth faltering early should reduce the risk of its severe consequences, including mortality, during this formative window of child development. Long-term investments and patience may be required, as it will take decades to eliminate the intergenerational factors limiting mothers’ size.

## Data Availability

The data that support the findings of this study are available from the Bill and Melinda Gates Foundation Knowledge Integration project upon reasonable request.

https://child-growth.github.io/causes/

https://github.com/child-growth/ki-longitudinal-growth

---

**Extended Data Figure 1.**
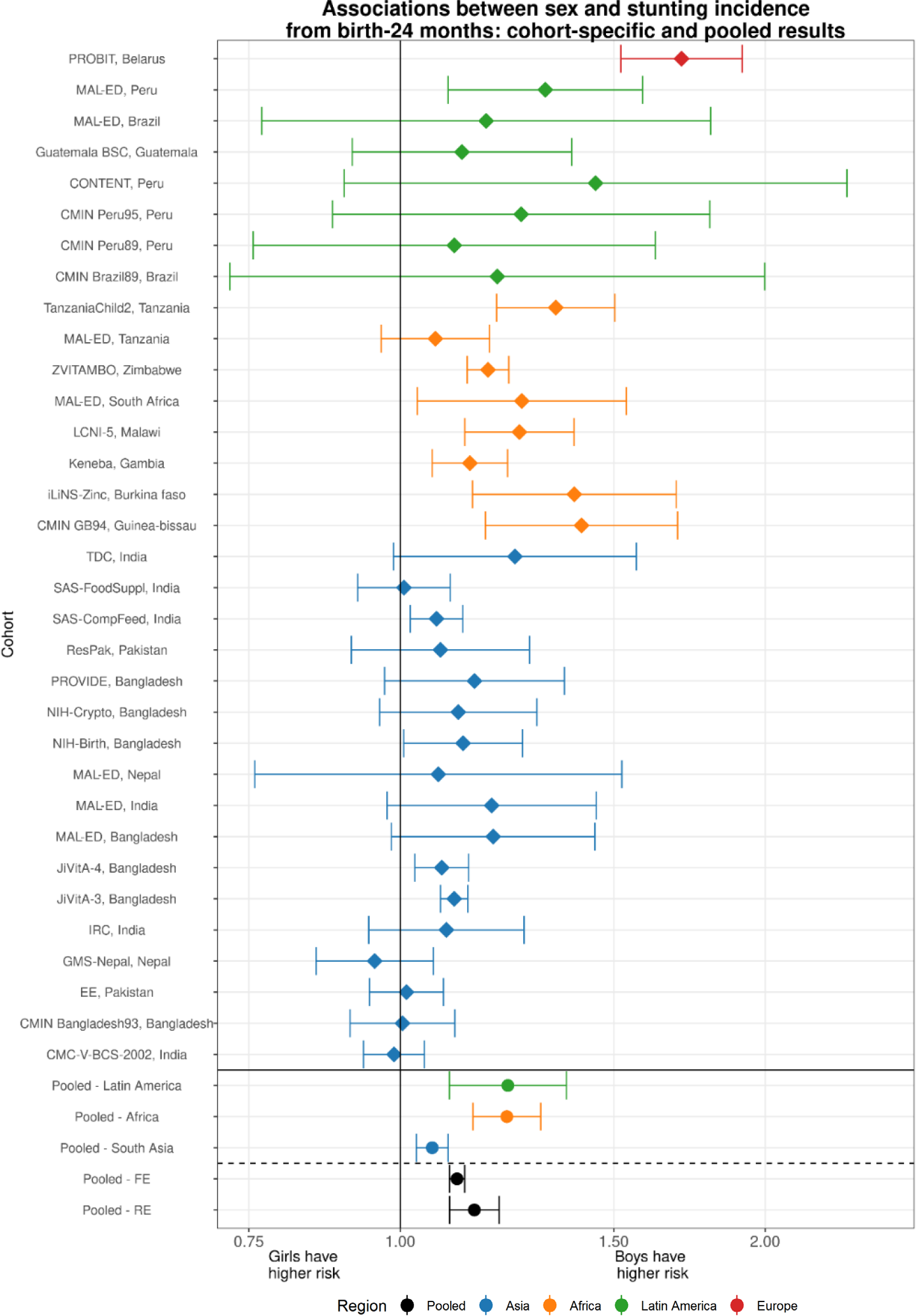
Example forest plot of cohort-specific and pooled parameter estimates. Cohort-specific estimates of the cumulative incidence ratio of stunting are plotted on each row, comparing the risk of any stunting from birth to 24 months among boys compared to a reference level of girls. Below the solid horizontal line are region-specific pooled measures of association, pooled using random-effects models. Below the dashed line are overall pooled measures of association, comparing pooling using random or fixed effects models. The primary results reported throughout the manuscript are overall (not region stratified) estimates pooled using random effects models.

**Extended Data Figure 2.**
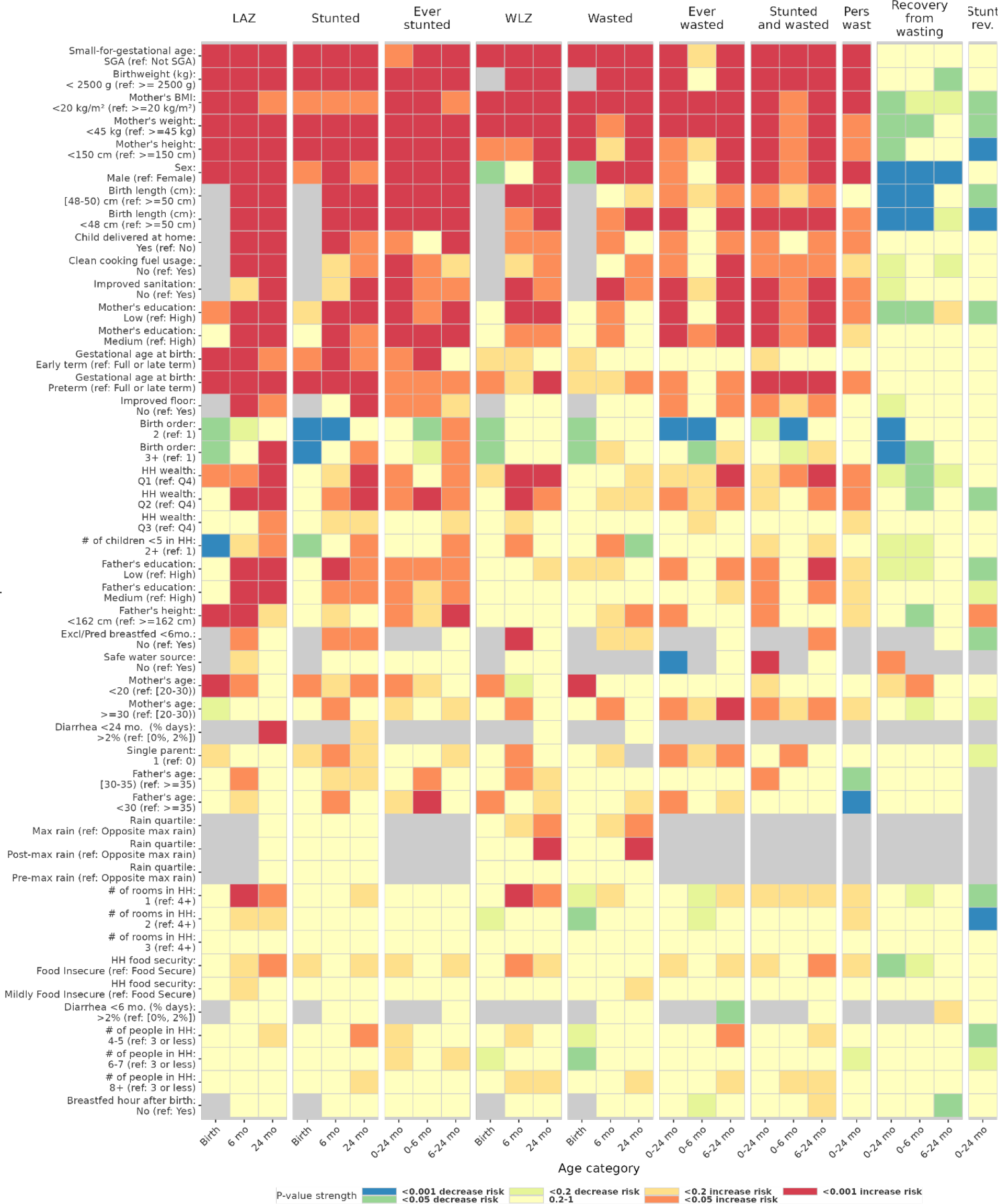
Heatmap of significance and direction across exposure-outcome combinations. The heatmap shows the significance and direction of estimates through the cell colors, separated across primary outcomes by child age. Red and orange cells are exposures where the outcome is estimated have an increased probability of occurring compared to the reference level (harmful exposures except for recovery outcomes), while blue and green cells are exposures associated with a decreased probability of the outcome (protective exposures except for recovery outcomes). The outcomes are labeled at the top of the columns, with each set of three columns the set of three ages analyzed for that outcome. Each row is a level of an exposure variable, with reference levels excluded. Rows are sorted top to bottom by increasing average p-value. Grey cells denote comparisons that were not estimated or could not be estimated because of data sparsity in the exposure-outcome combination. All point estimates and confidence intervals for exposure-outcome pairs with P-values plotted in this figure are viewable online at (https://child-growth.github.io/causes).

**Extended Data Figure 3.**
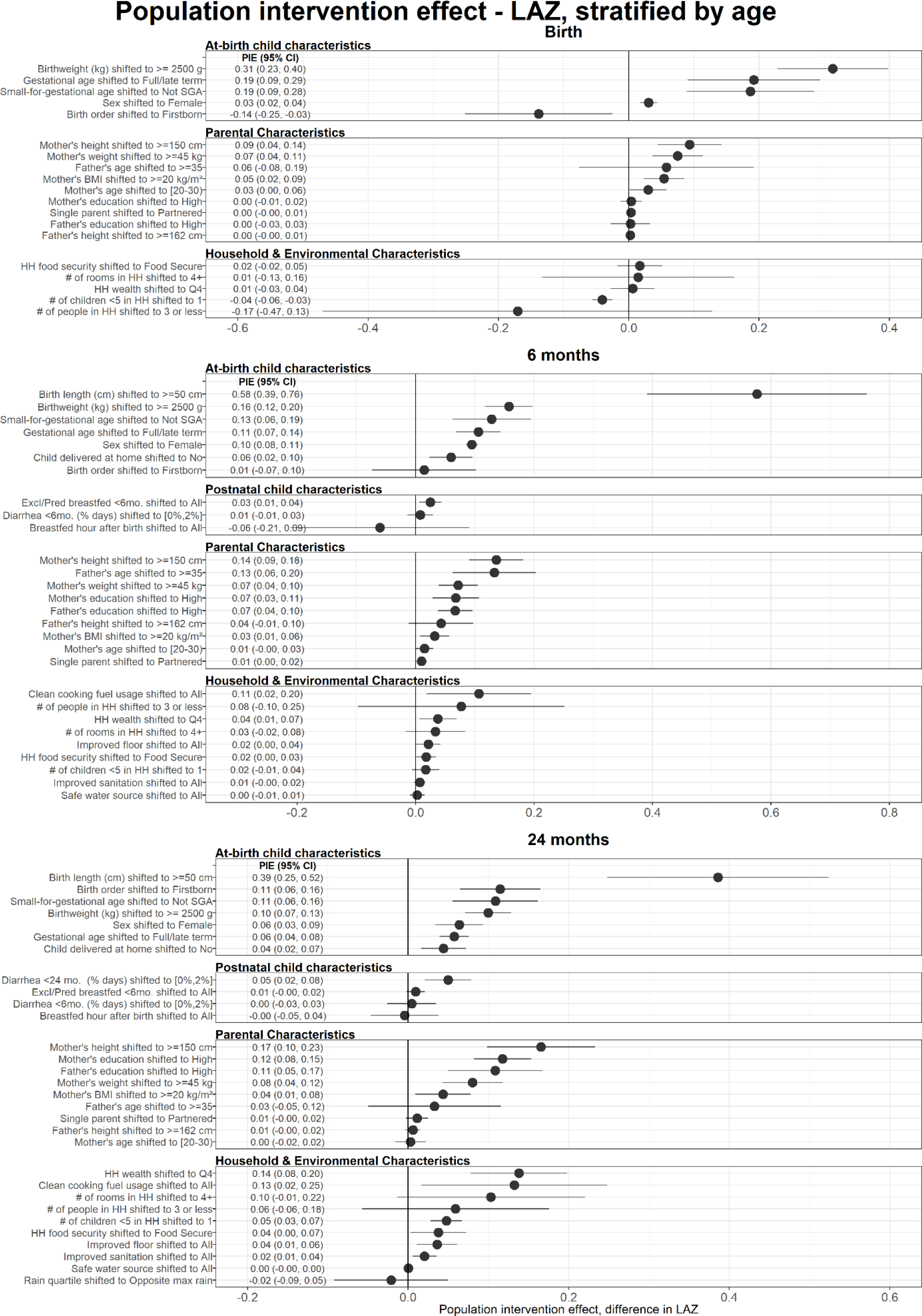
Age-stratified population intervention effects in length-for-age Z-scores. Exposures, rank ordered by population intervention effect on child LAZ, stratified by the age of the child at the time of anthropometry measurement. The population intervention effect is the expected difference in mean Z-score if all children had the reference level of the exposure rather than the observed exposure distribution. Reference levels are printed in the exposure label. Cohort-specific estimates were adjusted for all measured confounders using ensemble machine learning and TMLE, and then pooled using random effects (Methods). Estimates are shown only for exposures measured in at least 4 cohorts.

**Extended Data Figure 4.**
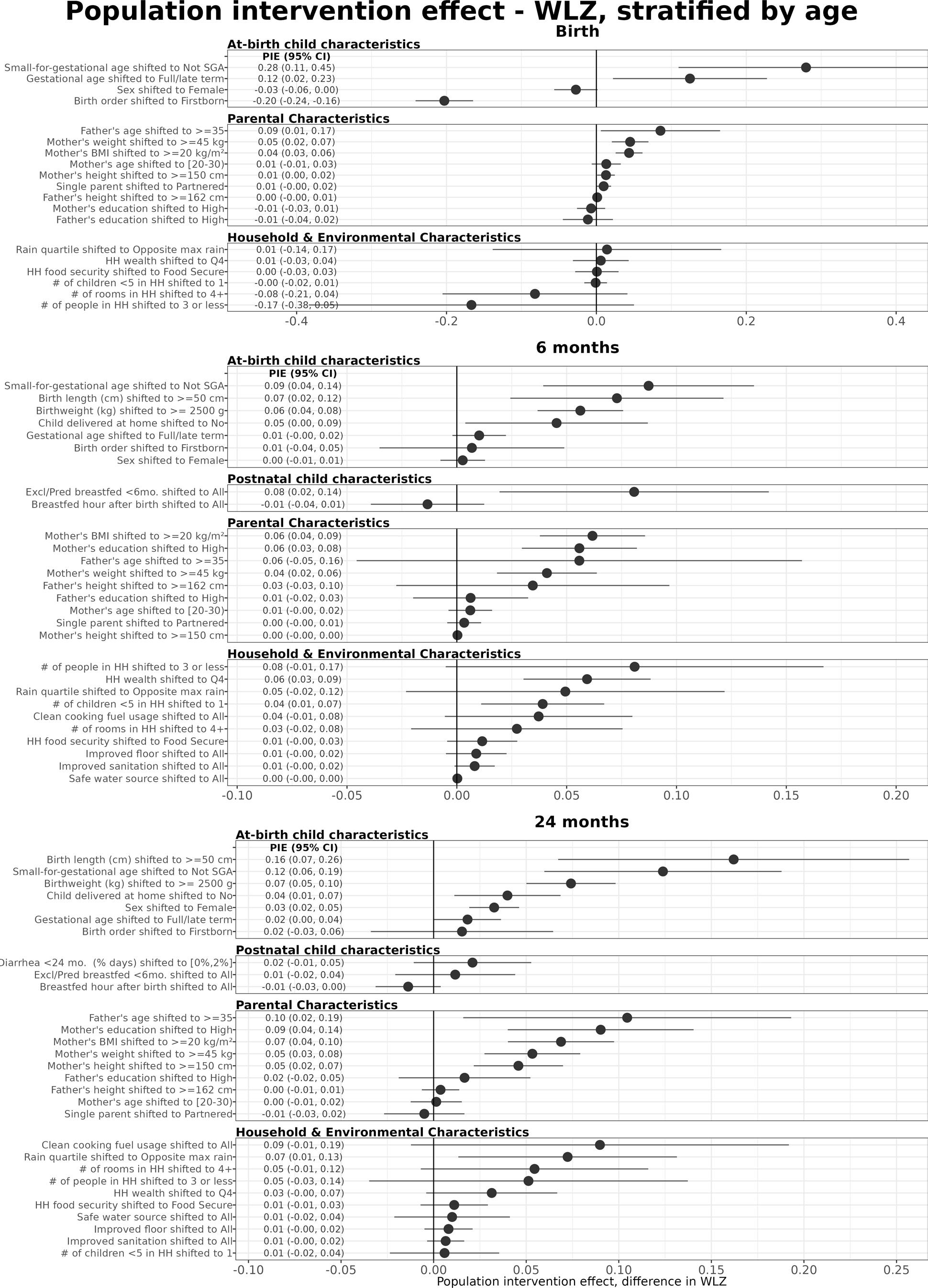
Age-stratified population intervention effects in weight-for-length Z-scores. Exposures, rank ordered by population intervention effects on child WLZ,, stratified by the age of the child at the time of anthropometry measurement. The population intervention effect is the expected difference in population mean Z-score if all children had the reference level of the exposure rather than the observed distribution. For all plots, reference levels are printed next to the name of the exposure. Cohort-specific estimates were adjusted for all measured confounders using ensemble machine learning and TMLE, and then pooled using random effects (Methods). Estimates are shown only for exposures measured in at least 4 cohorts.

**Extended Data Figure 5.**
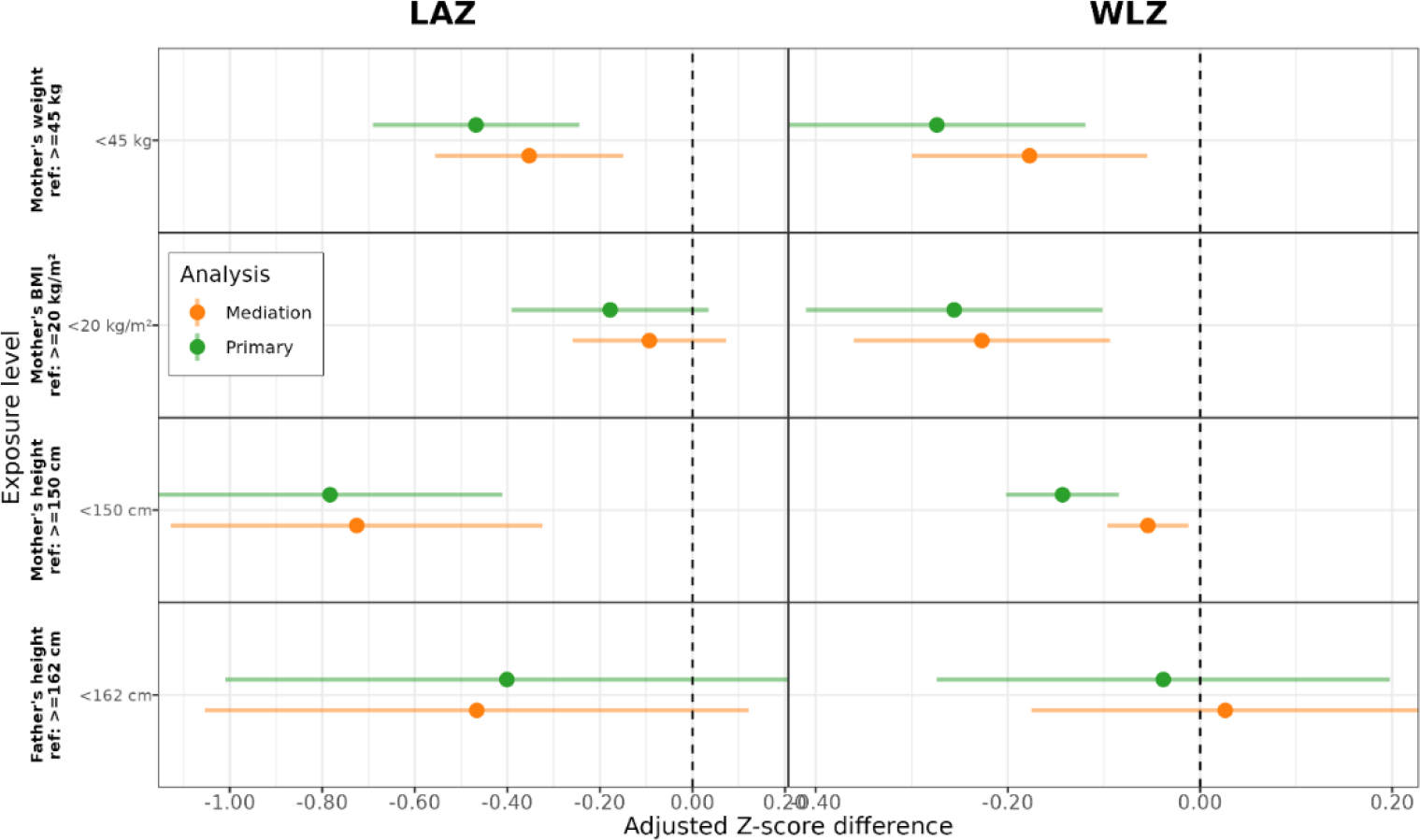
Mediation of parental anthropometry effects by birth size on child Z-scores at 24 months. Mediating effect of adjusting for birth anthropometry and at-birth characteristics on the estimated Z-score differences between levels of parental anthropometry. Primary estimates were adjusted for all other measured exposures not on the causal pathway, while the mediation analysis estimates were additionally adjusted for birthweight, birth length, gestational age at birth, birth order, small-for-gestational age status, and home vs. hospital delivery. Only estimates from cohorts measuring at least 3 of the 6 at-birth characteristics were used to estimate the pooled Z-score differences (n = 6 cohorts, 17,124 observations). Mediation estimates were slightly attenuated toward the null, and only in the case of maternal height and child WLZ were they statistically different from the primary analysis. These results imply that the causal pathway between parental anthropometry and growth faltering operates through its effect on birth size, but most of the effect is through other pathways.

**Extended Data Figure 6.**
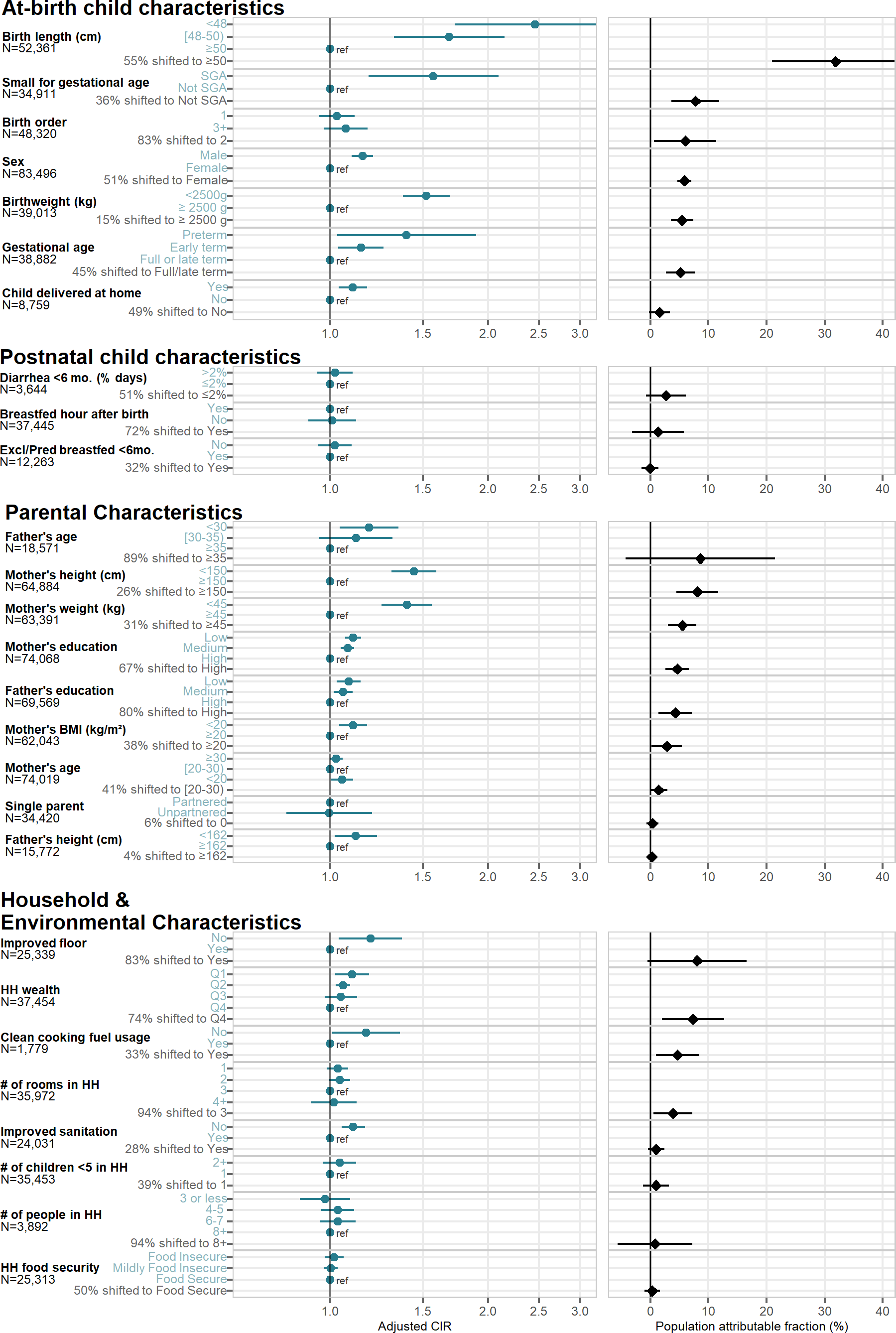
Rank-ordered associations between child, parental, and household characteristics and adjusted relative risks or population attributable fractions of stunting by age 24 months. Blue points in the left panel show adjusted cumulative incidence ratios (CIRs) between higher-risk exposure levels and reference levels, and black points in the right panel show population attributable fractions (PAFs), the estimated proportion of the risk in the whole population that would be removed if the exposure were set to its indicated reference level. The number of children that contributed to each analysis is listed by exposure. The colored Y-axis label is either the level of exposure contrasted against the reference level to estimate the CIR, or the percent of the population shifted to the lowest-risk level to estimate the PAF. For at-birth exposures, at-birth stunting and wasting were excluded to focus on incidence of new (postnatal) cases, and for postnatal exposures (breastfeeding practice and diarrheal disease), the cumulative incidence of stunting from 6-24 months was used. Cohort-specific estimates were adjusted for all measured confounders using ensemble machine learning and TMLE, and then pooled using random effects (Methods). Estimates are shown only for exposures measured in at least 4 studies.

**Extended Data Figure 7.**
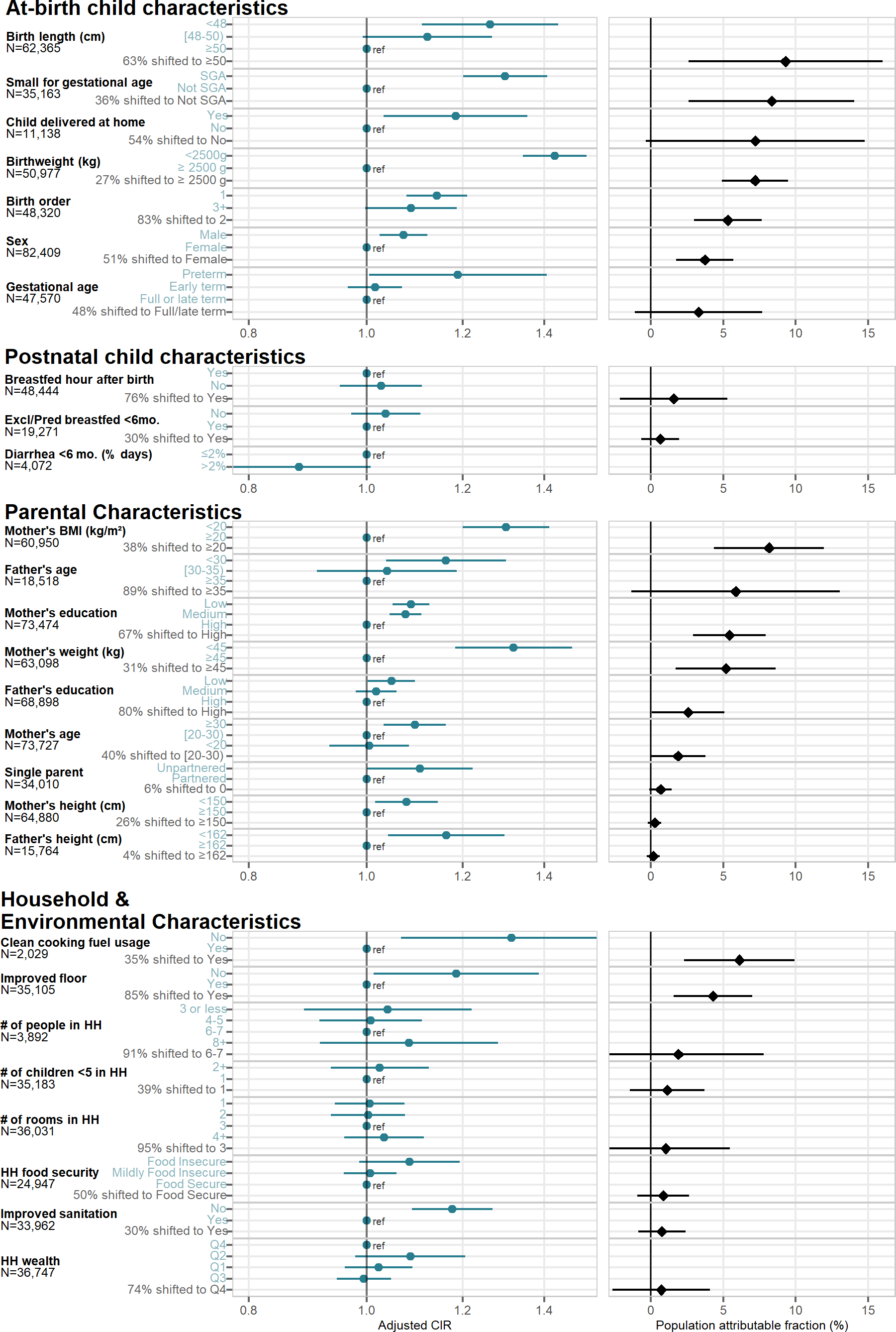
Rank-ordered associations between child, parental, and household characteristics and adjusted relative risks or population attributable fractions of wasting by age 24 months. Blue points in the left panel show adjusted cumulative incidence ratios (CIRs) between higher-risk exposure levels and reference levels, and black points in the right panel show population attributable fractions (PAFs), the estimated proportion of the risk in the whole population that would be removed if the exposure were set to its indicated reference level. The number of children that contributed to each analysis is listed by exposure. The colored Y-axis label is either the level of exposure contrasted against the reference level to estimate the CIR, or the percent of the population shifted to the lowest-risk level to estimate the PAF. For at-birth exposures, at-birth stunting and wasting were excluded, and for postnatal exposures (breastfeeding practice and diarrheal disease), the cumulative incidence of wasting from 6-24 months was used. Cohort-specific estimates were adjusted for all measured confounders using ensemble machine learning and TMLE, and then pooled using random effects (Methods). Estimates are shown only for exposures measured in at least 4 studies. The PAF for diarrhea under 6 months was not calculable or plotted due to the unexpected CIR <1 for estimated higher diarrheal disease burden.

**Extended Data Figure 8.**
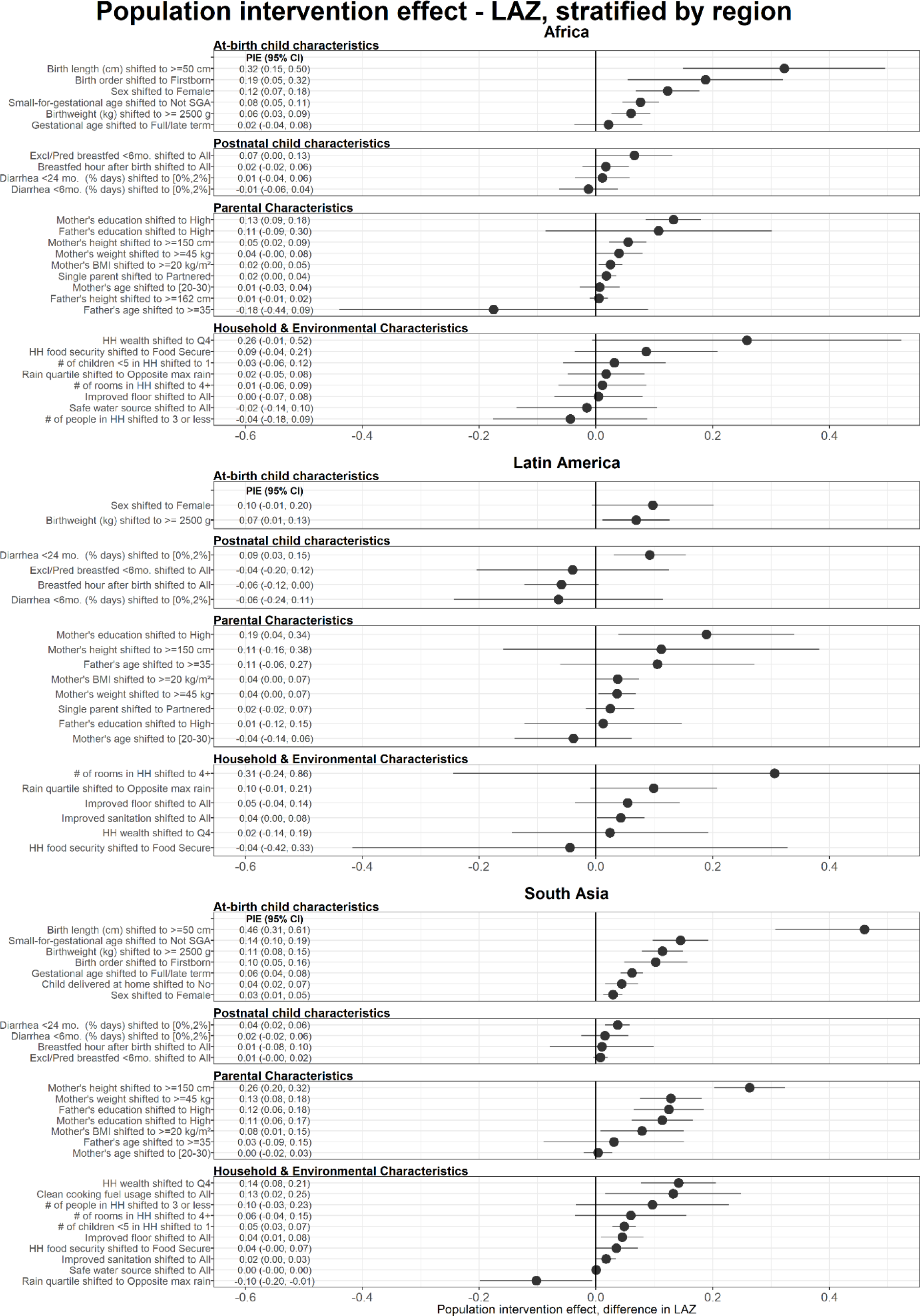
Regionally-stratified population intervention effects for length-for-age Z-scores at age 24 months. Exposures, rank ordered by population intervention effect on child length-for-age z-score (LAZ) at age 24 months, stratified by region. The population intervention effect is the expected difference in population mean Z-score if all children had the reference level of the exposure rather than the observed distribution. For all plots, reference levels are printed next to the name of the exposure. Cohort-specific estimates were adjusted for all measured confounders using ensemble machine learning and TMLE, and then pooled using random effects (Methods). Estimates are shown only for exposures measured in at least 4 cohorts.

**Extended Data Figure 9.**
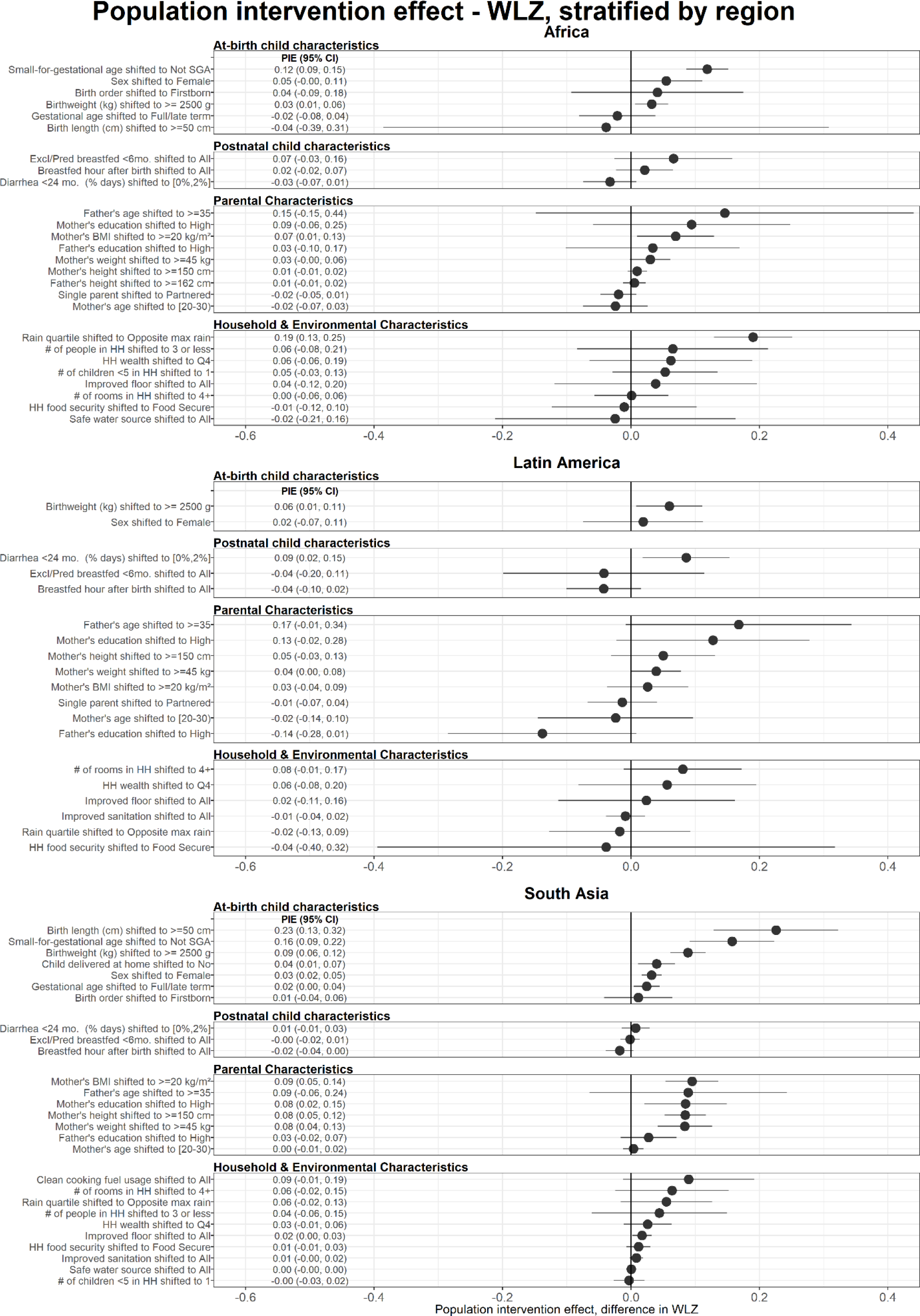
Regionally-stratified population intervention effects for weight-for-length Z-scores at age 24 months. Exposures, rank ordered by population attributable difference on child weight-for-length z-score (WLZ) at age 24 months, stratified by region. The population intervention effect is the expected difference in population mean Z-score if all children had the reference level of the exposure rather than the observed distribution. For all plots, reference levels are printed next to the name of the exposure. Cohort-specific estimates were adjusted for all measured confounders using ensemble machine learning and TMLE, and then pooled using random effects (Methods). Estimates are shown only for exposures measured in at least 4 cohorts.

**Extended Data Figure 10.**
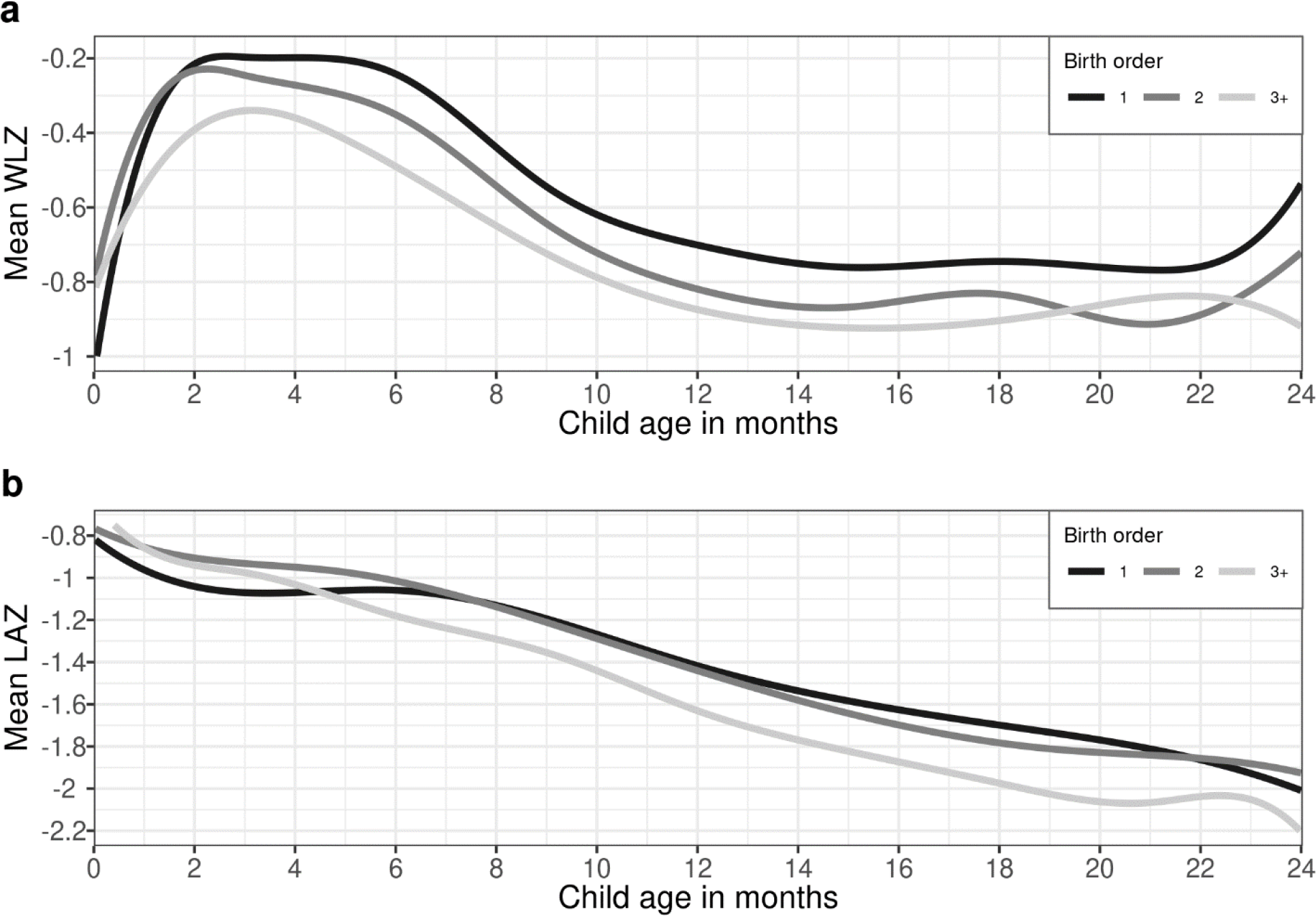
Child growth trajectories stratified by birth order (a) Child weight-for-length Z-score (WLZ) trajectories, stratified by categories of child birth order. (b) Child length-for-age Z-score (LAZ) trajectories, stratified by categories of child birth order. Details on the estimation of growth trajectories are in the Methods. Child growth trajectories stratified by categories of all risk factors are available online (https://child-growth.github.io/causes/rf-splines.html).

**Extended Data Figure 11.**
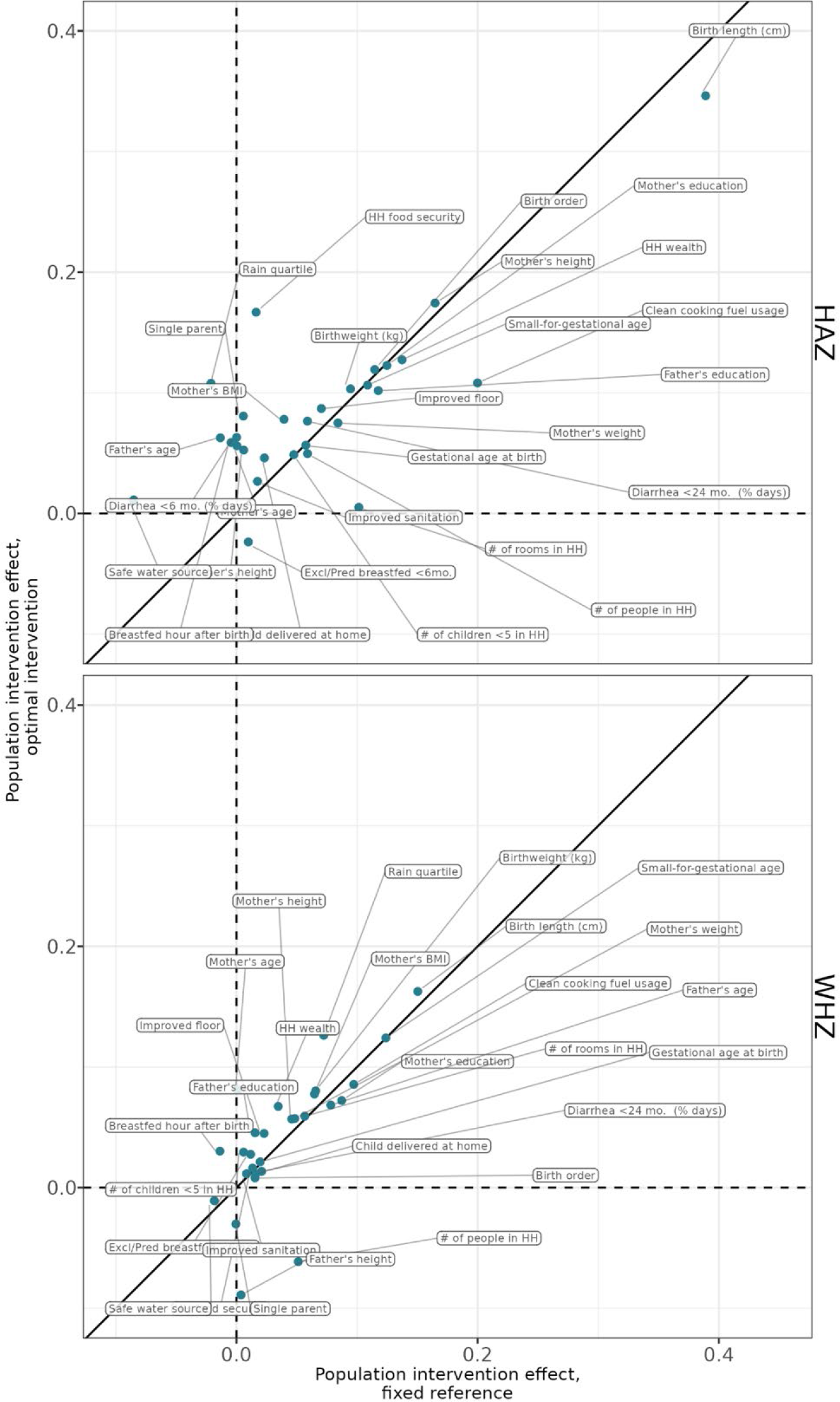
Comparing fixed-reference and optimal intervention estimates of the population intervention effect. Pooled population intervention effects on child LAZ and WHZ at 24 months, with the X-axis showing attributable differences using a fixed, and the Y-axis showing the optimal intervention attributable difference, where the level the exposure is shifted to can vary by child. Points are labeled with the specific risk factor. Estimates farther from the diagonal line have larger differences between the static and optimal intervention estimates. The optimal intervention attributable differences, which are not estimated with an a-priori specified low-risk reference level, were generally close to the static attributable differences, indicating that the chosen reference levels were the lowest risk strata in most or all children.

**Extended Data Figure 12.**
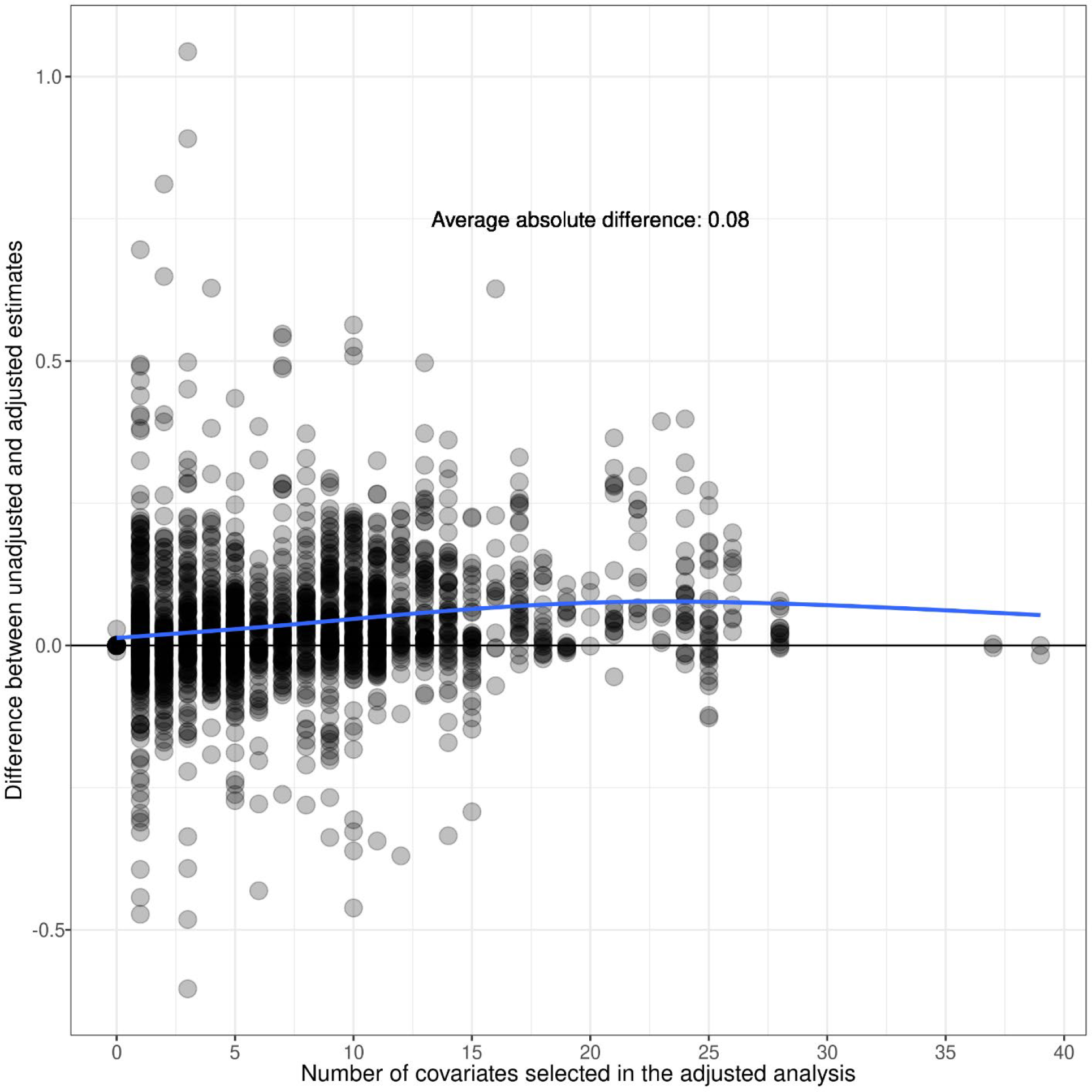
Difference between adjusted and unadjusted Z-score effects by number of selected adjustment variables. Points mark the difference in estimates unadjusted and adjusted estimates of the difference in average Z-scores between exposed and unexposed children across 33 cohorts, 30 exposures and length-for-age and weight-for-length Z-score outcomes included in the analysis. Different cohorts measured different sets of exposures, and a different number of adjustment covariates were chosen for each cohort-specific estimate based on outcome sparsity, so cohort-specific estimates adjust for different covariates and numbers of covariates. The plot shows no systematic bias between unadjusted and adjusted estimates based on number of covariates chosen. The blue line shows the average difference between adjusted estimates from unadjusted estimates, fitted using a cubic spline.

**Extended Data Figure 13.**
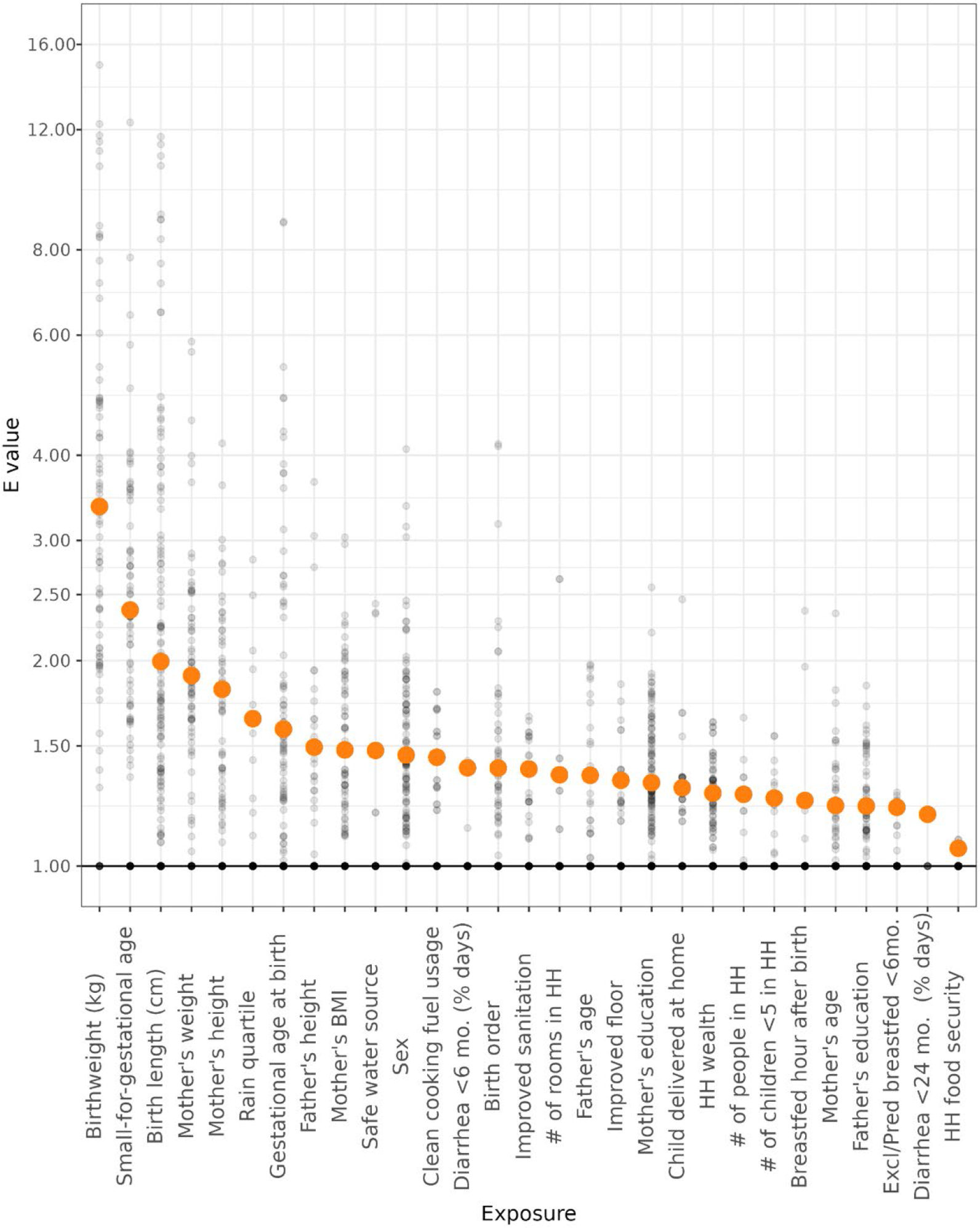
Assessing sensitivity of estimates to unmeasured confounding using E-values An E-value is the minimum strength of association in terms of relative risk that an unmeasured confounder would need to have with both the exposure and the outcome to explain away an estimated exposure–outcome association.^1^ Orange points mark the E-values for the pooled estimates of relative risk for each exposure. Grey points are cohort-specific E-values for each exposure-outcome relationship. Non-significant pooled estimates have points plotted at 1.0. Orange points are median E-values among statistically significant estimates for each exposure. As an example, an unmeasured confounder would on average need to almost double the risk of both the exposure and the outcome to explain away observed significant associations for the birth length exposure.

**Extended data table 1.**
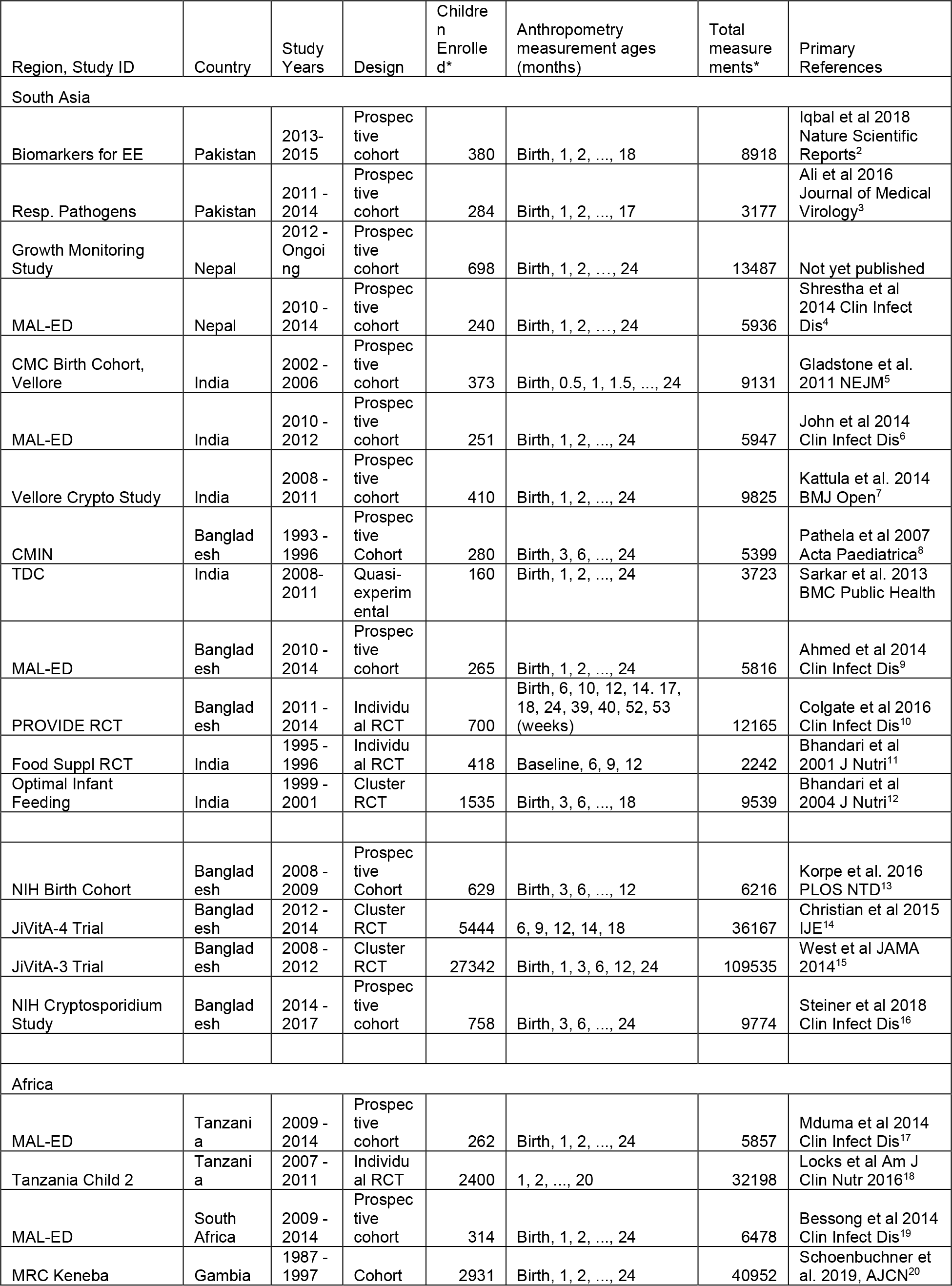
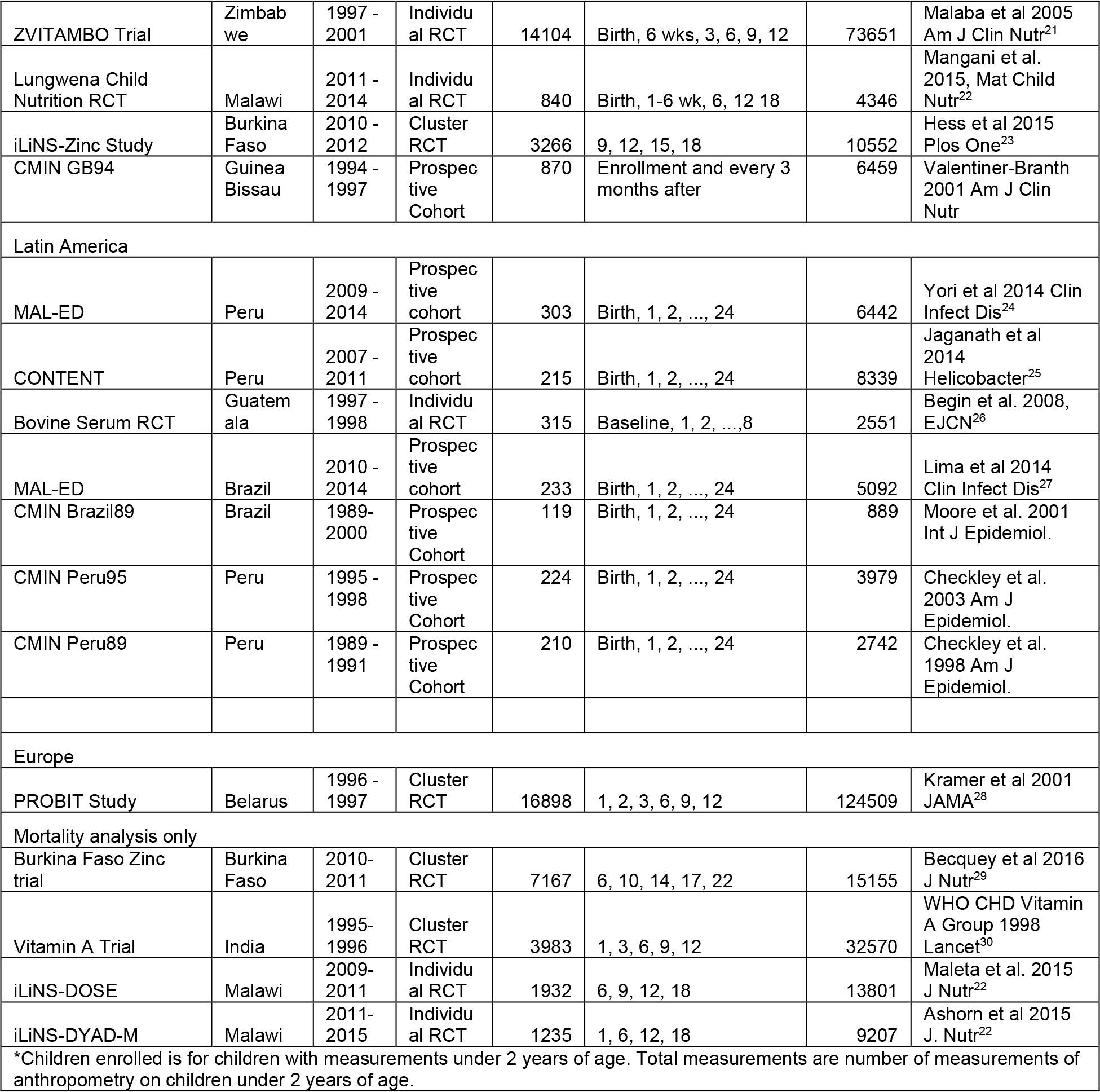

**Extended data table 2.**
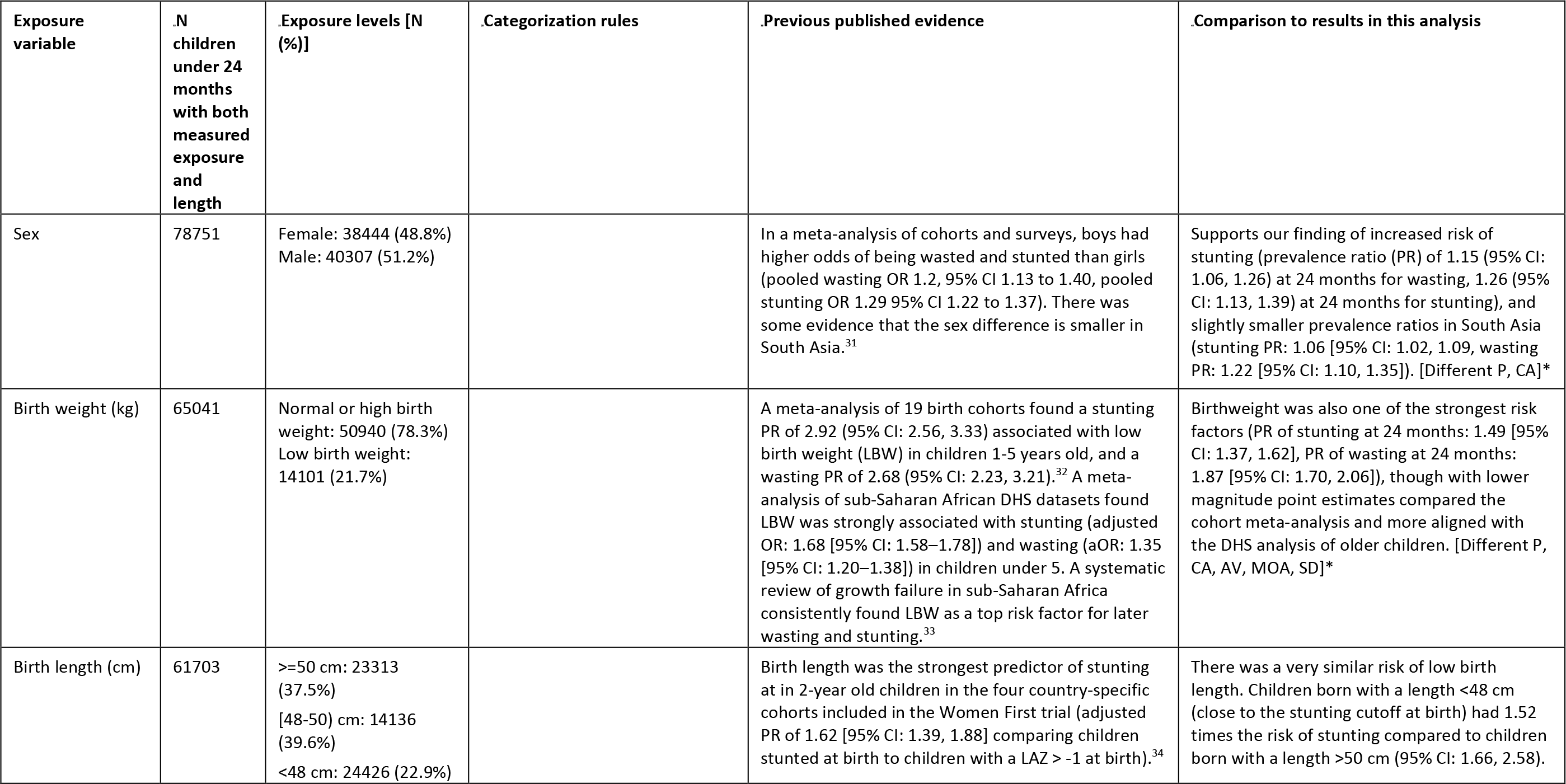
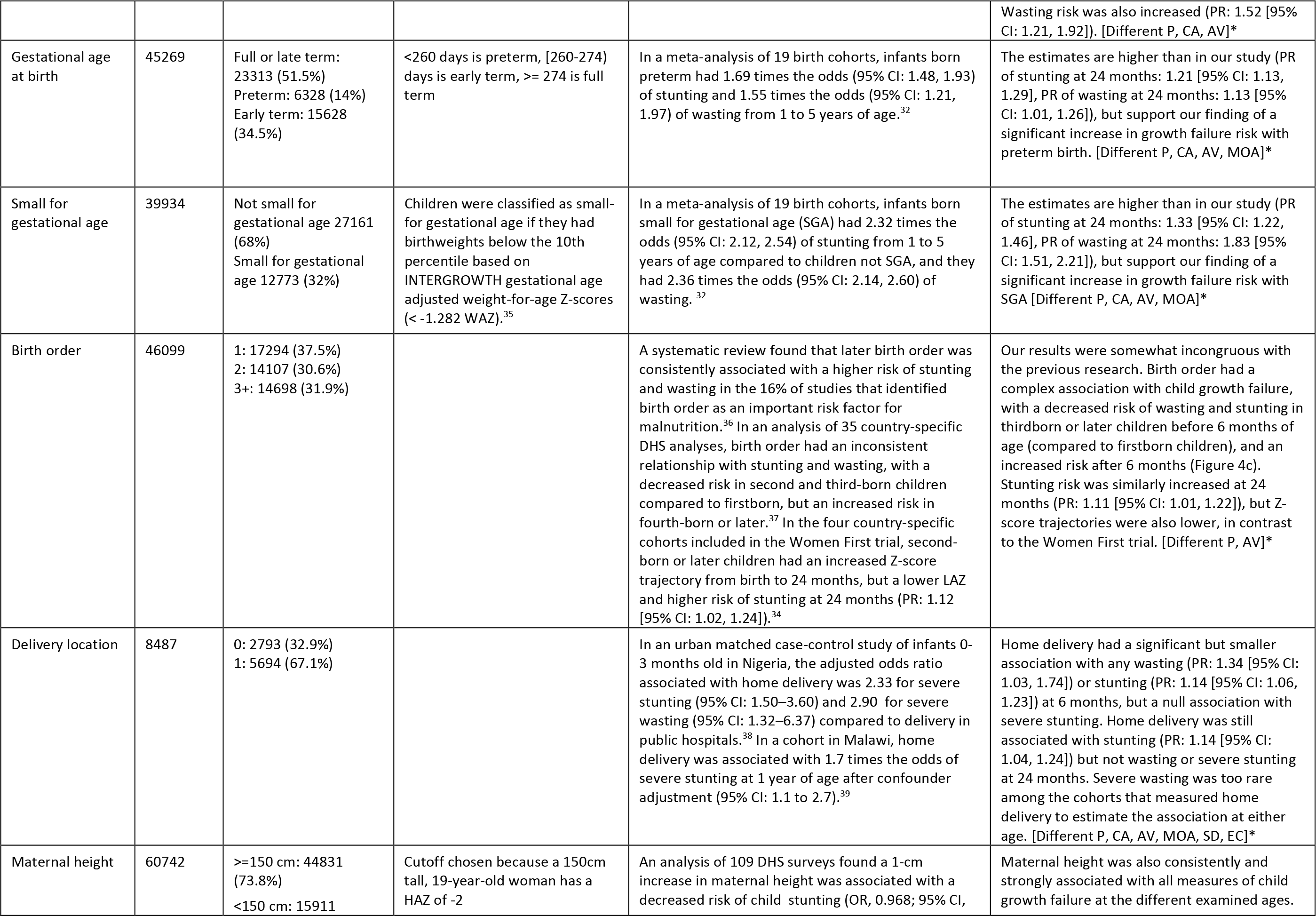
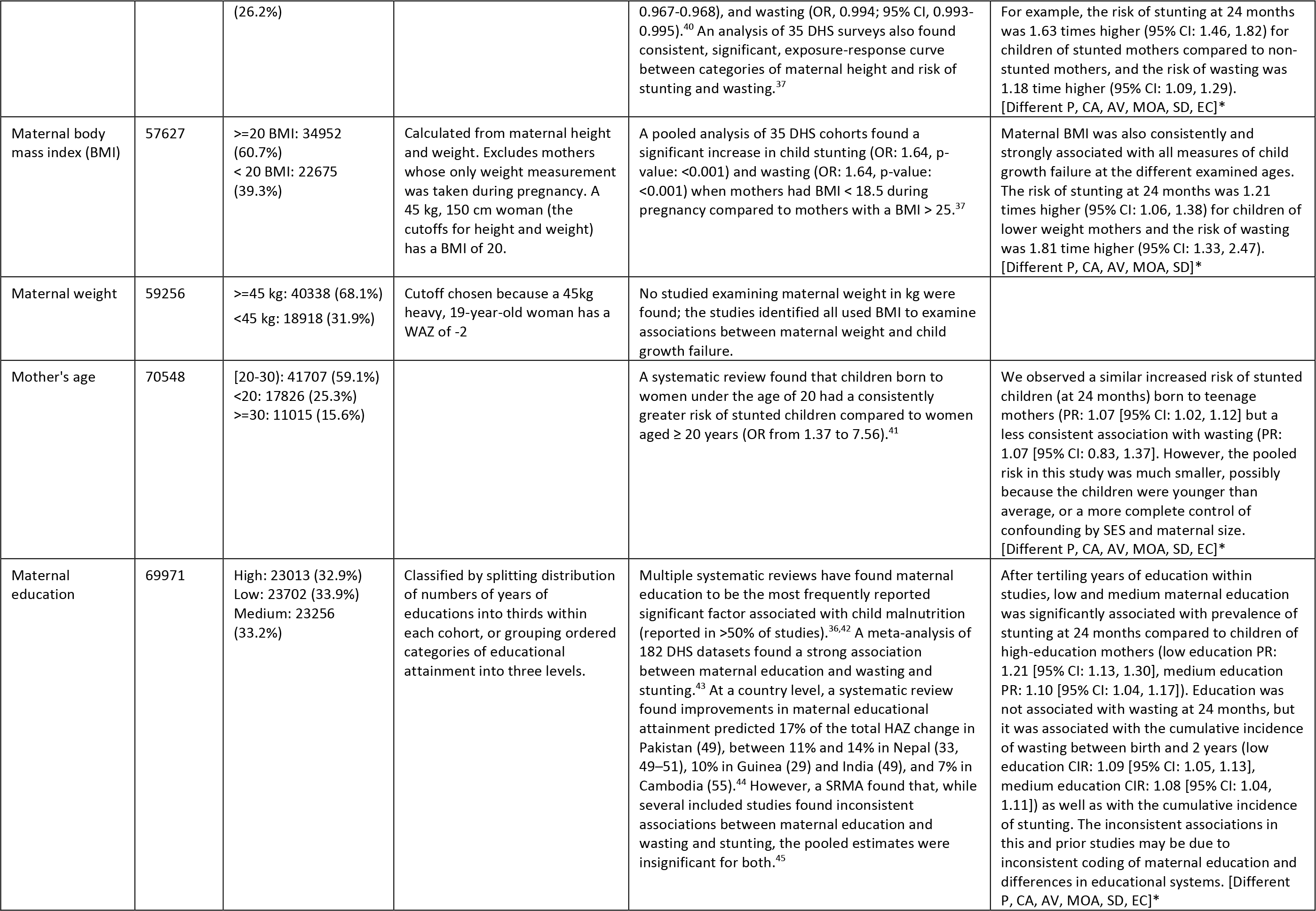
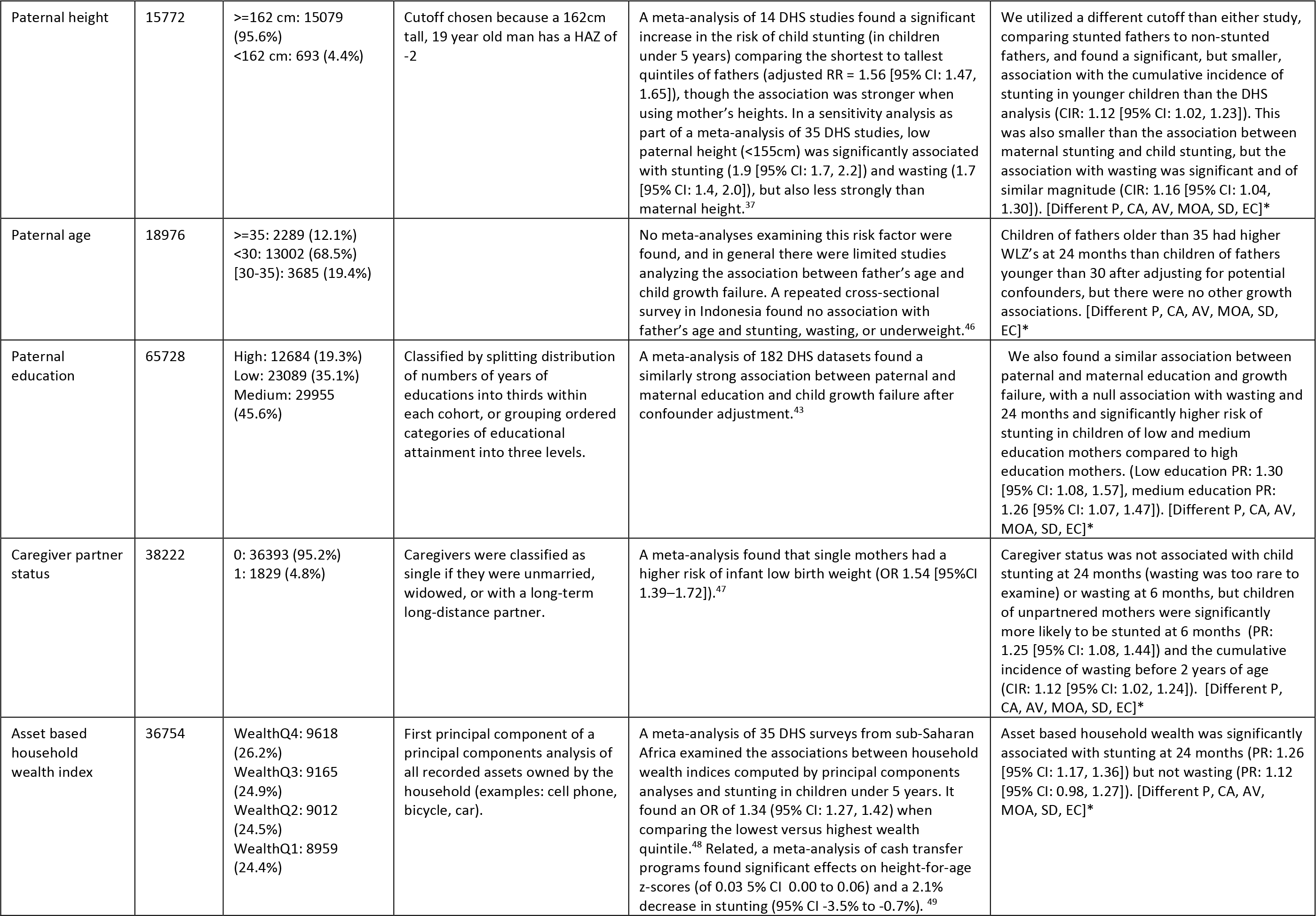
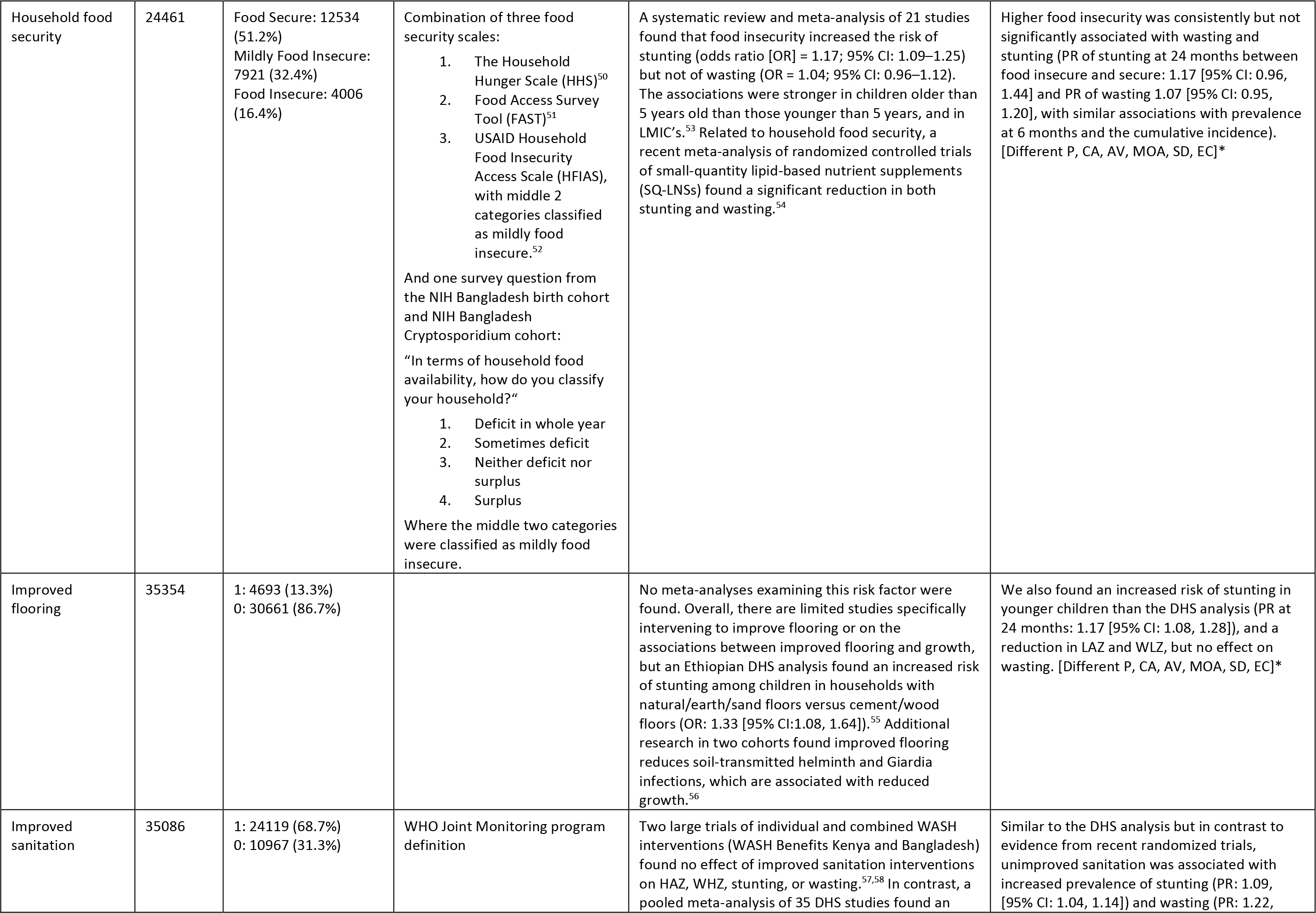
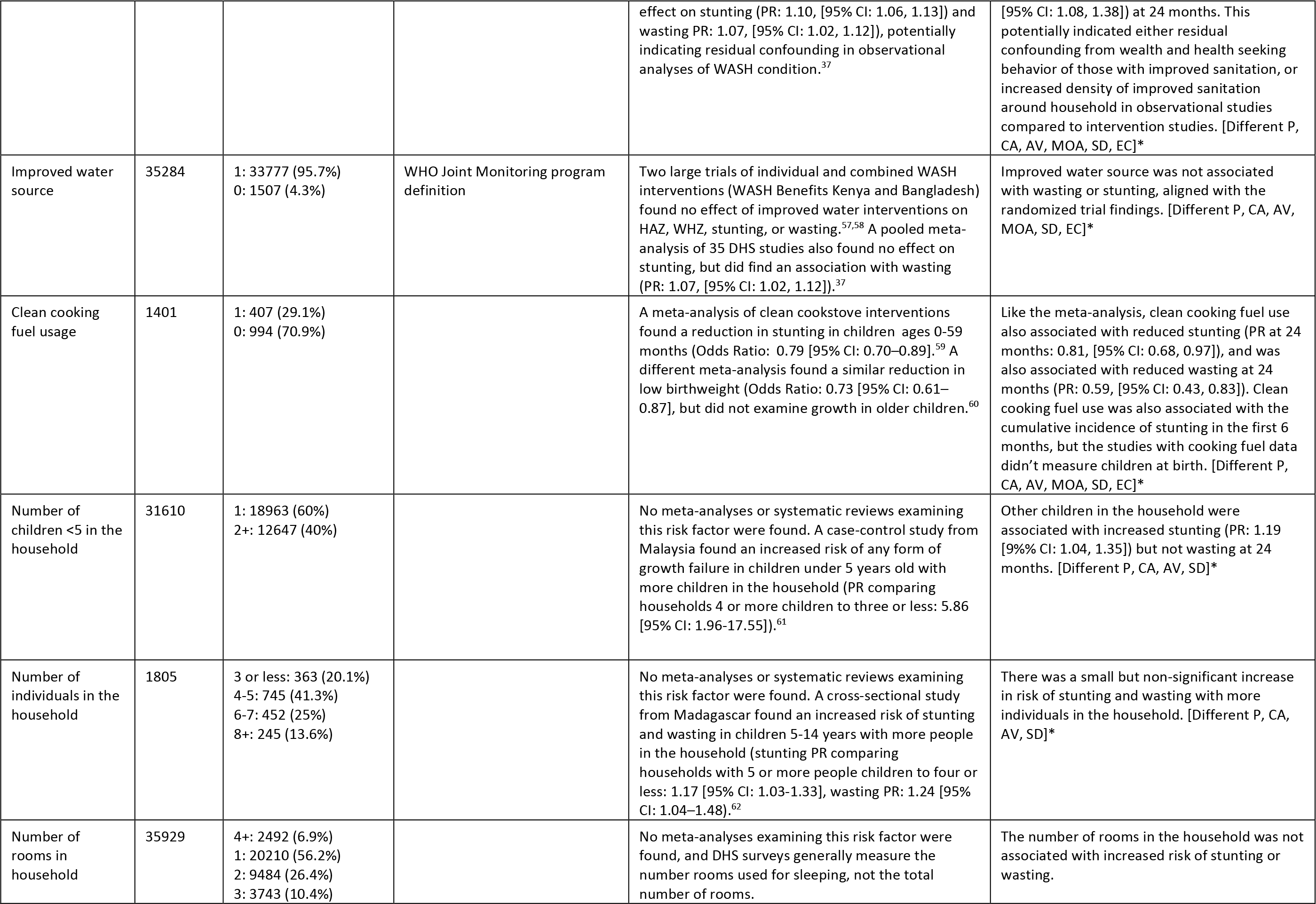
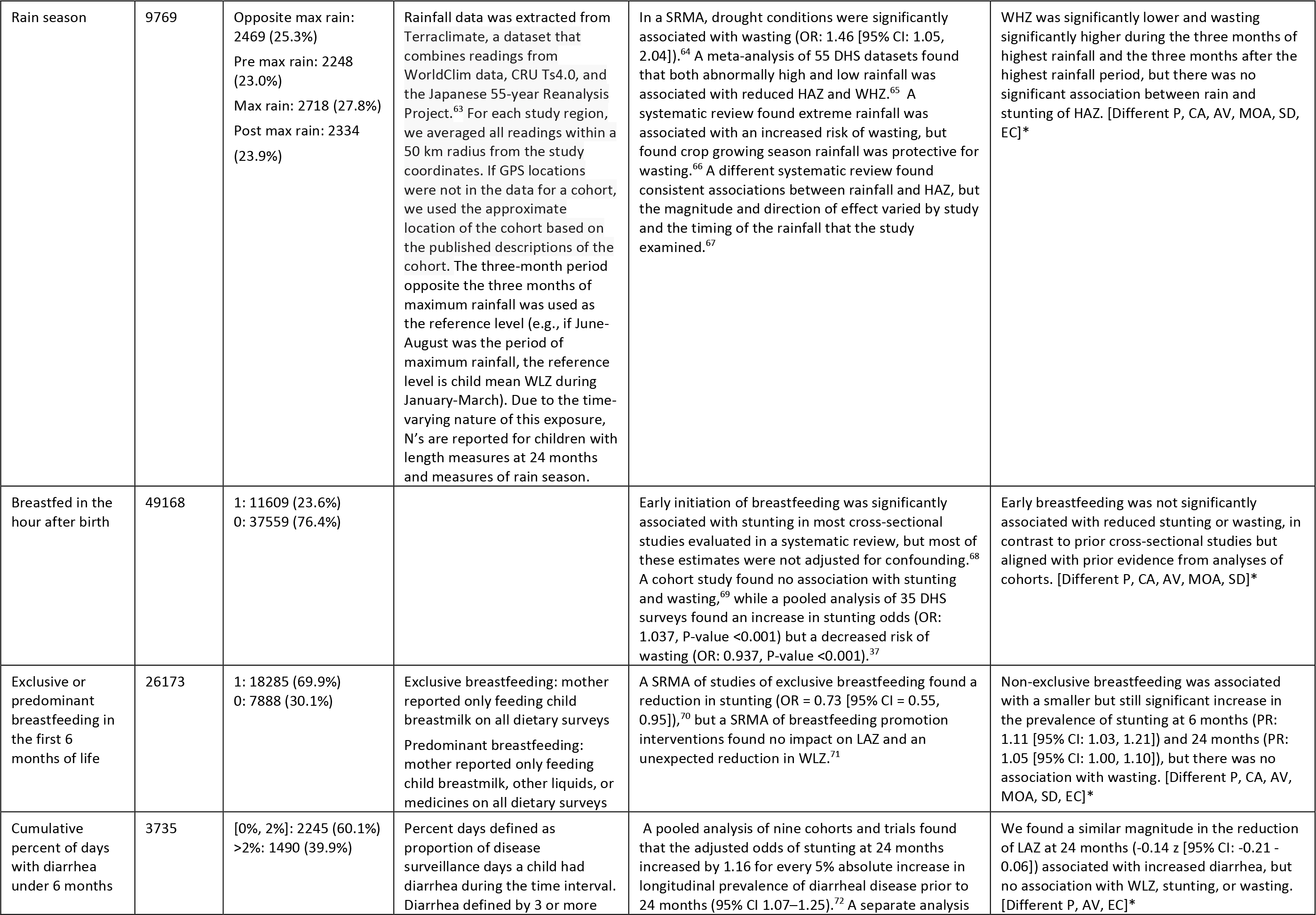
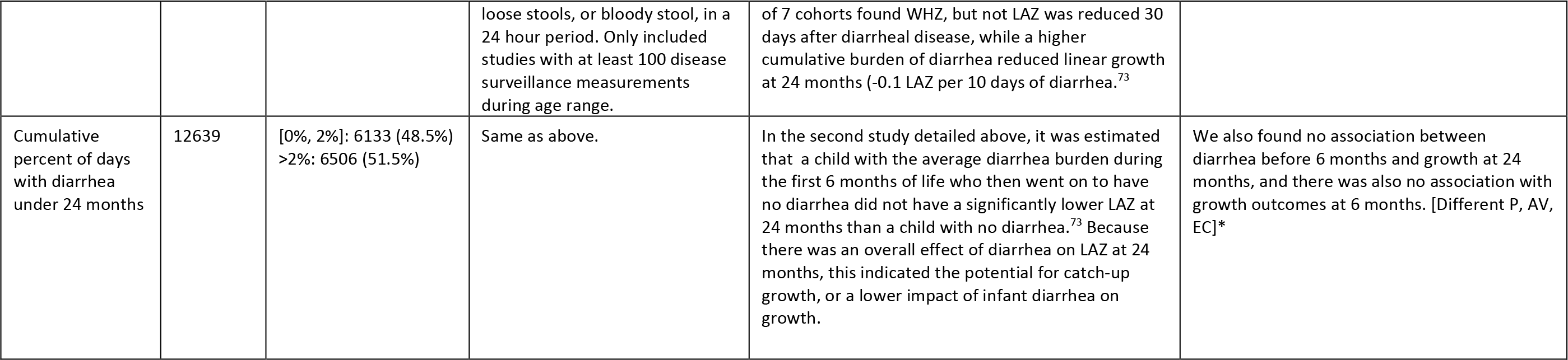
All exposures included in the analysis, as well as the categories the exposures were classified into across all cohorts, categorization rules, the total number of children, the percentage of children in each category, select evidence from prior literature, and comparisons to our results. We selected the exposures of interest based on variables present in multiple cohorts that met our inclusion criteria, were found to be important determinants of stunting and wasting in prior literature, and could be harmonized across cohorts for pooled analyses. Where possible, we cite findings from recent randomized controlled trials and systematic reviews. All referenced results from this manuscript are available here: https://child-growth.github.io/causes/RR-forest.html. *Bracketed codes at the end of each cell in the “Comparison to results in this analysis” indicate limitations to comparisons with previous evidence due to differences in: P=population, CA=child age, AV=adjustment variables used in the analysis, MOA=measure of association, SD=study design, EC=exposure classification.

**Extended data table 3.** Under 1-year country-specific mortality rate is from UNICEF (https://data.unicef.org/country), and is higher than the cohort-specific under 2-year mortality rate for all cohorts used in the mortality analysis.

| Study | Country | Number of deaths under 2 | Under 2 mortality rate in cohort (%) | Infant (Under 1) mortality rate in cohort (%) | Infant (Under 1) mortality country rate (UNICEF) |
| --- | --- | --- | --- | --- | --- |
| Burkina Faso Zn | Burkina Faso | 39 | 0.54 | 0.42 | 5.4 |
| iLiNS-DOSE | Malawi | 53 | 2.74 | 1.92 | 3.1 |
| iLiNS-DYAD-M | Malawi | 54 | 4.37 | 3.48 | 3.1 |
| JiVitA-3 | Bangladesh | 934 | 3.41 | 2.85 | 2.6 |
| JiVitA-4 | Bangladesh | 49 | 0.9 | 0.39 | 2.6 |
| Keneba | The Gambia | 65 | 2.22 | 1.52 | 3.6 |
| VITAMIN-A | India | 108 | 2.70 | 2.7 | 2.8 |
| ZVITAMBO | Zimbabwe | 1113 | 7.89 | 6.57 | 3.8 |

## Materials and Methods

### 1. Study designs and inclusion criteria

We included all longitudinal observational studies and randomized trials available through the *ki* project on April 1, 2018 that met five inclusion criteria: 1) conducted in low- or middle-income countries; 2) enrolled children between birth and age 24 months and measured their length and weight repeatedly over time; 3) did not restrict enrollment to acutely ill children; 4) enrolled children with a median year of birth after 1990; 5) collected anthropometry measurements at least quarterly. We included all children under 24 months of age, assuming months were 30.4167 days, and we considered a child’s first measure recorded by age 7 days as their anthropometry at birth. Four additional studies with high-quality mortality information that measured children at least every 6 months were included in the mortality analyses (The Burkina Faso Zinc trial, The Vitamin-A trial in India, and the iLiNS-DOSE and iLiNS-DYAD-M trials in Malawi).

### 2. Statistical analysis

Analyses were conducted in R version 4.0.5. All pooled, regional, and cohort-specific results, results for secondary outcomes, and sensitivity analyses are available online at (https://child-growth.github.io/causes).

### 3. Outcome definitions

We calculated length-for-age Z-scores (LAZ), weight-for-age Z-scores (WAZ), and weight-for-length Z-scores (WLZ) using WHO 2006 growth standards.^1^ We used the medians of triplicate measurements of heights and weights of children from pre-2006 cohorts to re-calculate Z-scores to the 2006 standard. We dropped 1,190 (0.2%) unrealistic measurements of LAZ (>+6 or <–6 Z), 1,330 (0.2%) measurements of WAZ (> 5 or < –6 Z), and 1,670 (0.3%) measurements of WLZ (>+5 or –5 Z), consistent with WHO recommendations.^2^ See Benjamin-Chung (2020) for more details on cohort inclusion and assessment of anthropometry measurement quality.^3^ We also calculated the difference in linear and ponderal growth velocities over three-month periods. We calculated the change in LAZ, WAZ, length in centimeters, and weight in kilograms within 3-month age intervals, including measurements within a two-week window around each age in months to account for variation in the age at each length measurement.

We defined stunting as LAZ < –2, severe stunting as LAZ < –3, underweight as WAZ < –2, severe underweight as WAZ < –3, wasting as WLZ < –2, severe wasting as WLZ < –3, concurrent stunting and wasting as LAZ < –2 and WLZ < –2. Children with ≥ 50% of WLZ measurements < –2 and at least 4 measurements over a defined age range were classified as persistently wasted (e.g., birth to 24 months, median interval between measurements: 80 days, IQR: 62-93). Children were assumed to never recover from stunting episodes, but children were classified as recovered from wasting episodes (and at risk for a new episode of wasting) if their measured WLZ was ≥ –2 for at least 60 days (details in Mertens et. al (2020)).^4^ Stunting reversal was defined as children stunted under 3 months whose final two measurements before 24 months were non-stunted. Child mortality was all-cause and was restricted to children who died after birth and before age 24 months. For child morbidity outcomes (Figure 4c), concurrent wasting and stunting prevalences at age 18 months were estimated using the anthropometry measurement taken closest to age 18 months, and within 17-19 months of age, while persistent wasting was estimated from child measurements between 6 and 24 months of age. We chose 18 months to calculate concurrent wasting and stunting because it maximized the number of child observations at later ages when concurrent wasting and stunting was most prevalent, and used ages 6- 24 months to define persistent wasting to maximize the number of anthropometry measurements taken after the early growth faltering exposure measurements.^4^

### 4. Estimating relationships between child, parental, and household exposures and measures of growth faltering

#### 4.1 Exposure definitions

We selected the exposures of interest based on variables present in multiple cohorts that met our inclusion criteria, were found to be important predictors of stunting and wasting in prior literature and could be harmonized across cohorts for pooled analyses. Extended Data Table 2 lists all exposures included in the analysis, as well as exposure categories used across cohorts, and the total number of children in each category. For parental education and asset-based household wealth, we categorized to levels relative to the distribution of educational attainment within each cohort. Continuous biological characteristics (gestational age, birth weight, birth height, parental weight, parental height, parental age) were classified based on a common distribution, pooling data across cohorts. Our rationale was that the meaning of socio- economic variables is culturally context-dependent, whereas biological variables should have a more universal meaning.

#### 4.2 Risk set definition

For exposures that occur or exist before birth, we considered the child at risk of incident outcomes at birth. Therefore, we classified children who were born stunted (or wasted) as incident episodes of stunting (or wasting) when estimating the relationship between household characteristics, paternal characteristics, and child characteristics like gestational age, sex, birth order, and birth location.

For postnatal exposures (e.g., breastfeeding practices, WASH characteristics, birth weight), we excluded episodes of stunting or wasting that occurred at birth. Children who were born wasted could enter the risk set for postnatal exposures if they recovered from wasting during the study period (see Mertens et al. 2020 for details).^4^ This restriction ensured that for postnatal exposures, the analysis only included postnatal, incident episodes. Children born or enrolled wasted were included in the risk set for the outcome of recovery from wasting within 90 days for all exposures (prenatal and postnatal).

#### 4.3 Estimating differences in outcomes across categories of exposures

We estimated measures of association between exposures and growth faltering outcomes by comparing outcomes across categories of exposures in four ways:

<u>Mean difference</u> of the comparison levels of the exposure on LAZ, WLZ at birth, 6 months, and 24 months. The Z-scores used were the measures taken closest to the age of interest and within one month of the age of interest, except for Z-scores at birth which only included a child’s first measure recorded by age 7 days. We also calculated mean differences in LAZ, WAZ, weight, and length velocities.

<u>Prevalence ratios (PR)</u> between comparison levels of the exposure, compared to the reference level at birth, 6 months, and 24 months. Prevalence was estimated using anthropometry measurements closest to the age of interest and within one month of the age of interest, except for prevalence at birth which only included measures taken on the day of birth.

Cumulative incidence ratios (CIR) between comparison levels of the exposure, compared to the reference level, for the incident onset of outcomes between birth and 24 months, 6-24 months, and birth-6 months.

<u>Mean Z-scores by continuous age, stratified by levels of exposures,</u> from birth to 24 months were fit within individual cohorts using cubic splines with the bandwidth chosen to minimize the median Akaike information criterion across cohorts.^5^ We estimated splines separately for each exposure category. We pooled spline curves across cohorts into a single prediction, offset by mean Z-scores at one year, using random effects models.^6^

#### 4.4 Estimating population attributable parameters

We estimated three measures of the population-level effect of exposures on growth faltering outcomes:

<u>Population intervention impact</u> (PIE), a generalization of population attributable risk, was defined as the change in population mean Z-score if the entire population’s exposure was set to an ideal reference level. For each exposure, we chose reference levels based on prior literature or as the category with the highest mean LAZ or WLZ across cohorts.

<u>Population attributable fraction</u> (PAF) was defined as the proportional reduction in cumulative incidence if the entire population’s exposure was set to an ideal low risk reference level. We estimated the PAF for the prevalence of stunting and wasting at birth, 6, and 24 months and cumulative incidence of stunting and wasting from birth to 24 months, 6-24 months, and from birth to 6 months. For each exposure, we chose the reference level as the category with the lowest risk of stunting or wasting.

<u>Optimal individualized intervention impact</u> We employed a variable importance measure (VIM) methodology to estimate the impact of an optimal individualized intervention on an exposure.^7^ The optimal intervention on an exposure was determined through estimating individualized treatment regimes, which give an individual-specific rule for the lowest-risk level of exposure based on individuals’ measured covariates. The covariates used to estimate the low-risk level are the same as those used for the adjustment documented in section 6 below. The impact of the optimal individualized intervention is derived from the VIM, which is the predicted change in the population-mean outcome from the observed outcome if every child’s exposure was shifted to the optimal level. This differs from the PIE and PAF parameters in that we did not specify the reference level; moreover, the reference level could vary across participants.

PIE and PAF parameters assume a causal relationship between exposure and outcome. For some exposures, we considered attributable effects to have a pragmatic interpretation — they represent a summary estimate of relative importance that combines the exposure’s strength of association and its prevalence in the population.^8^ Comparisons between optimal intervention estimates and PIE estimates are shown in Extended Data Fig 11.

### 5. Estimation approach

#### Estimation of cohort-specific effects

For each exposure, we used the directed acyclic graph (DAG) framework to identify potential confounders from the broader set of exposures used in the analysis.^9^ We did not adjust for characteristics that were assumed to be intermediate on the causal path between any exposure and the outcome, because while controlling for mediators may help adjust for unmeasured confounders in some conditions, it can also lead to collider bias.^10, 11^ Detailed lists of adjustment covariates used for each analysis are available online (https://child-growth.github.io/causes/dags.html). Confounders were not measured in every cohort, so there could be residual confounding in cohort-specific estimates.

Analyses used a complete case approach that only included children with non-missing exposure and outcome measurements. For additional covariates in adjusted analyses, we used the following approach to impute missing covariate values.^12^ Within each cohort, if there was <50% missingness in a covariate, we imputed missing measurements as the median (continuous variables) or mode (categorical variables) among all children, and analyses included an indicator variable for missingness in the adjustment set. Covariates with >50% missingness were excluded from the potential adjustment set.

When calculating the median for imputation, we used children as independent units rather than measurements so that children with more frequent measurements were not over-represented.

Unadjusted PRs and CIRs between the reference level of each exposure and comparison levels were estimated using logistic regressions.^13^ Unadjusted mean differences for continuous outcomes were estimated using linear regressions.

To flexibly adjust for potential confounders and reduce the risk of model misspecification, we estimated adjusted PRs, CIRs, and mean differences using targeted maximum likelihood estimation (TMLE), a two-stage estimation strategy that incorporates state-of-the-art machine learning algorithms (super learner) while still providing valid statistical inference. ^14, 15^ The effects of covariate adjustment on estimates compared to unadjusted estimates is shown in Extended Data Fig 12, and E-values, summary measures of the strength of unmeasured confounding needed to explain away observed significant associations, are plotted in Extended Data Fig 13.^16^ The super learner is an ensemble machine learning method that uses cross-validation to select a weighted combination of predictions from a library of algorithms.^17^ We included in the library simple means, generalized linear models, LASSO penalized regressions,^18^ generalized additive models,^19^ and gradient boosting machines.^20^ The super learner was fit to maximize the 10-fold cross-validated area under the receiver operator curve (AUC) for binomial outcomes, and minimize the 10-fold cross-validated mean-squared error (MSE) for continuous outcomes. That is, the super learner was fit using 9/10 of the data, while the AUC/MSE was calculated on the remaining 1/10 of the data. Each fold of the data was held out in turn and the cross-validated performance measure was calculated as the average of the performance measures across the ten folds. This approach is practically appealing and robust in finite samples, since this cross-validation procedure utilizes unseen sample data to measure the estimator’s performance. Also, the super learner is asymptotically optimal in the sense that it is guaranteed to outperform the best possible algorithm included in the library as sample size grows. The initial estimator obtained via super learner is subsequently updated to yield an efficient double-robust semi-parametric substitution estimator of the parameter of interest.^14^ To estimate the R^2^ of models including multiple exposures, we fit super learner models, without the targeted learning step, and within each cohort measuring the exposures. We then pooled cohort-specific R^2^ estimates using fixed effects models.

We estimated influence curve-based, clustered standard errors to account for repeated measures in the analyses of recovery from wasting or progression to severe wasting. We assumed that the children were the independent units of analysis unless the original study had a clustered design, in which case the unit of independence in the original study were used as the unit of clustering. We used clusters as the unit of independence for the iLiNS-Zinc, Jivita-3, Jivita-4, Probit, and SAS Complementary Feeding trials. We estimated 95% confidence intervals for incidence using the normal approximation.

Mortality analyses estimated hazard ratios using Cox proportional hazards models with a child’s age in days as the timescale, adjusting for potential confounders, with the growth faltering exposure status updated at each follow-up that preceded death or censoring by age 24 months. Growth faltering exposures included moderate (between –2 Z and –3 Z) wasting, stunting, and underweight, severe (below –3 Z) wasting, stunting, and underweight, and combinations of concurrent wasting, stunting, and underweight. Growth faltering categories were mutually exclusive within moderate or severe classifications, so children were classified as only wasted, only stunted, or only underweight, or some combination of these categories. We estimated the hazard ratio associated with different anthropometric measures of CGF in separate analyses, considering each as an exposure in turn with the reference group defined as children without the deficit. For children who did not die, we defined their censoring date as the administrative end of follow-up in their cohort, or age 24 months (730 days), whichever occurred first. Because mortality was a rare outcome, estimates are adjusted only for child sex and trial treatment arm. To avoid reverse causality, we did not include child growth measures occurring within 7 days of death. Extended Data Table 3 lists the cohorts used in the mortality analysis, the number of deaths in each cohort, and a comparison to country-level infant mortality rates.

#### Data sparsity

We did not estimate relative risks between a higher level of exposure and the reference group if there were 5 or fewer cases in either stratum. In such cases, we still estimated relative risks between other exposure strata and the reference strata if those strata were not sparse. For rare outcomes, we only included one covariate for every 10 observations in the sparsest combination of the exposure and outcome, choosing covariates based on ranked deviance ratios.

### 6. Pooling parameters

We pooled adjusted estimates from individual cohorts using random effects models, fit using restricted maximum likelihood estimation. The pooling methods are detailed in Benjamin-Chung (2020).^1^ All parameters were pooled directly using the cohort-specific estimates of the same parameter, except for population attributable fractions. Pooled PAFs were calculated from random-effects pooled population intervention impacts (PIEs), and pooled outcome prevalence in the population using the following formulas:^21^

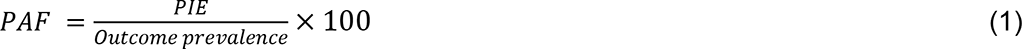

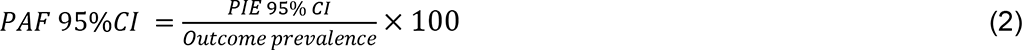

For PAFs of exposures on the cumulative incidence of wasting and stunting, the pooled cumulative incidence was substituted for the outcome prevalence in the above equations. We used this method instead of direct pooling of PAFs because, unlike PAFs, PIEs are unbounded with symmetrical confidence intervals.

For figures 3a-c, mean trajectories estimated using cubic splines in individual studies and then curves were pooled using random effects.^6^ Curves estimated from all anthropometry measurements of children taken from birth to 24 months of age within studies that measured the measure of maternal anthropometry.

### 7. Sensitivity analyses

We compared estimates pooled using random effects models, which are more conservative in the presence of heterogeneity across studies, with estimates pooled using fixed effects, and we compared adjusted estimates with estimates unadjusted for potential confounders. We estimated associations between growth faltering and mortality at different ages, after dropping the trials measuring children less frequently than quarterly, and using TMLE instead of Cox proportional hazard models, and we plotted Kaplan Meier curves of child mortality, stratified by measures of early growth faltering. We also conducted a sensitivity analysis on methods of pooling splines of child growth trajectories, stratified by maternal anthropometry. We re-estimated the attributable differences of exposures on WLZ and LAZ at 24 months, dropping the PROBIT trial, the only European study. Results from secondary outcomes and sensitivity analyses are viewable online at https://child-growth.github.io/causes.

### Data and code availability

The data that support the findings of this study are available from the Bill & Melinda Gates Foundation Knowledge Integration project upon reasonable request. Replication scripts for this analysis are available here: https://osf.io/9xyqv/.

## Acknowledgments

This research was financially supported by a global development grant (OPP1165144) from the Bill & Melinda Gates Foundation to the University of California, Berkeley, CA, USA. We would also like to thank the following collaborators on the included cohorts and trials for their contributions to study planning, data collection, and analysis: Muhammad Sharif, Sajjad Kerio, Ms. Urosa, Ms. Alveen, Shahneel Hussain, Vikas Paudel (Mother and Infant Research Activities), Anthony Costello (University College London), Noel Rouamba, Jean-Bosco Ouédraogo, Leah Prince, Stephen A Vosti, Benjamin Torun, Lindsey M Locks, Christine M McDonald, Roland Kupka, Ronald J Bosch, Rodrick Kisenge, Said Aboud, Molin Wang, Azaduzzaman, Abu Ahmed Shamim, Rezaul Haque, Rolf Klemm, Sucheta Mehra, Maithilee Mitra, Kerry Schulze, Sunita Taneja, Brinda Nayyar, Vandana Suri, Poonam Khokhar, Brinda Nayyar, Poonam Khokhar, Jon E Rohde, Tivendra Kumar, Jose Martines, Maharaj K Bhan, and all other members of the study staffs and field teams. We would also like to thank all study participants and their families for their important contributions. We are grateful to the LCNI5 and iLiNS research teams, participants and people of Lungwena, Namwera, Mangochi and Malindi, our research assistants for their positive attitude, support, and help in all stages of the studies.

## Author contributions

Conceptualization: A.M., J.B., J.M.C., K.H.B., P.C., B.F.A

Funding Acquisition: J.M.C., A.E.H., M.J.V., B.F.A.

Data curation: A.M., J.B., J.C., O.S., W.C., A.N., N.N.P., W.J., E.C, E.O.C., S.D., N.H., I.M., H.L., R.H., V.S., J.H., T.N.

Formal analyses: A.M., J.B., J.C., O.S., W.C., A.N., N.N.P., W.J., E.C, E.O.C., S.D., N.H., I.M., H.L., V.S., B.F.A

Methodology: A.M., J.B., J.M.C, J.C., O.S., N.H., I.M., A.E.H., M.J.V.,K.H.B., P.C., B.F.A.

Visualization: A.M.,J.B., A.N., N.N.P., S.D., A.S., E.C, J.C., R.H., K.H.B., P.C., B.F.A.

Writing – Original Draft Preparation: A.M., J.B., B.F.A.

Writing – Review & Editing: A.M., J.B., J.M.C., K.H.B., P.C., B.F.A., *ki* Child Growth Consortium members

## Competing interest declaration

Thea Norman is an employee of the Bill & Melinda Gates Foundation (BMGF). Kenneth H Brown and Parul Christian are former employees of BMGF. Jeremy Coyle, Vishak Subramoney, Ryan Hafen, and Jonas Häggström work as research contractors funded by the BMGF.

## Additional information

Supplementary Information is available for this paper at https://child-growth.github.io/causes.

Correspondence and requests for materials should be addressed to Andrew Mertens and Benjamin F. Arnold.

## Notes

### Author Declarations

The Committee for Protection of Human Subjects at UC Berkeley determined that this analysis was exempt from review.

### Summary of Updates

Updated primary figures to show PIE and risk difference estimates alongside each other, added small-for-gestational age as a risk factor, included a scoping literature review of prior evidence, and updated writing throughout.

